# First-in-human immunoPET imaging of COVID-19 convalescent patients using dynamic total-body PET and a CD8-targeted minibody

**DOI:** 10.1101/2023.03.14.23287121

**Authors:** Negar Omidvari, Terry Jones, Pat M Price, April L Ferre, Jacqueline Lu, Yasser G Abdelhafez, Fatma Sen, Stuart H Cohen, Kristin Schmiedehausen, Ramsey D Badawi, Barbara L Shacklett, Ian Wilson, Simon R Cherry

## Abstract

With the majority of CD8^+^ T cells residing and functioning in tissue, not blood, developing noninvasive methods for *in vivo* quantification of their biodistribution and kinetics in humans offers the means for studying their key role in adaptive immune response and memory. This study is the first report on using positron emission tomography (PET) dynamic imaging and compartmental kinetic modeling for *in vivo* measurement of whole-body biodistribution of CD8^+^ T cells in human subjects. For this, a ^89^Zr-labeled minibody with high affinity for human CD8 (^89^Zr-Df-Crefmirlimab) was used with total-body PET in healthy subjects (N=3) and in COVID-19 convalescent patients (N=5). The high detection sensitivity, total-body coverage, and the use of dynamic scans enabled the study of kinetics simultaneously in spleen, bone marrow, liver, lungs, thymus, lymph nodes, and tonsils, at reduced radiation doses compared to prior studies. Analysis and modeling of the kinetics was consistent with T cell trafficking effects expected from immunobiology of lymphoid organs, suggesting early uptake in spleen and bone marrow followed by redistribution and delayed increasing uptake in lymph nodes, tonsils, and thymus. Tissue-to-blood ratios from the first 7 h of CD8-targeted imaging showed significantly higher values in the bone marrow of COVID-19 patients compared to controls, with an increasing trend between 2 and 6 months post-infection, consistent with net influx rates obtained by kinetic modeling and flow cytometry analysis of peripheral blood samples. These results provide the platform for using dynamic PET scans and kinetic modelling to study total-body immunological response and memory.

## INTRODUCTION

Understanding the adaptive immune response to viral infections and subsequent immunological memory is critical for development of vaccines and therapeutic options. Studying the immune response in humans has been conventionally focused on peripheral blood assays, particularly in longitudinal studies, due to complexity and invasive nature of tissue sampling approaches. However, the majority of immune cells involved in the adaptive immune response and immunological memory reside and function in tissue, particularly in lymphoid organs such as bone marrow, spleen, tonsils, and lymph nodes *(1,2)*. CD8^+^ T cells are one of the key players in cell-mediated immune response against viral infections and there has been a growing interest in studying the critical role of CD8^+^ T cell trafficking and preferential residence of CD8^+^ memory T cells in certain niches, such as bone marrow, in immunological memory *(3,4,5,6)*.

The recent pandemic caused by the severe acute respiratory syndrome coronavirus 2 (SARS-CoV-2) emphasized the need to better understand the role of adaptive immunity in viral infections and there has been particular interest in the role of T cell response to the coronavirus disease 2019 (COVID-19) *(7,8)*. Immunological memory to SARS-CoV-2 infection has been extensively characterized in blood *(9,10,11,12)*. In particular, SARS-CoV-2 specific CD8^+^ memory T cell response appears to persist for at least 8 months in blood, with a declining trend observed between 1-8 months post infection *(9,10)*. Furthermore, examination of SARS-CoV-2 seropositive organ donors has shown SARS-CoV-2–specific CD8^+^ T cell memory in bone marrow, spleen, lung, and lymph nodes for up to 6 months after infection *(13)*.

A non-invasive method capable of quantifying T cell density and trafficking rates in tissue at a systems level for the whole body could enable longitudinal studies in patients with viral infections and in the healthy populations, leading to better understanding of the adaptive immune response and immunological memory. The case for developing such a method for researching COVID-19 infection would include the acute and recovery phases, pre-existing immunity *(14)*, asymptomatic response *(15)*, and herd immunity *(16)*. These could be extended to studies of susceptibility to COVID-19 in association with age *(17)*, genetic factors *(18)*, gender *(19)*, children *(20)*, and obesity *(21)*. Being able to non-invasively study T cell involvement in peripheral effects *(22)*, long COVID *(23)*, vaccine efficacy *(24)*, and therapeutic interventions *(25)* would also suggest potentially fruitful areas for whole-body COVID-19 research.

As a proof of concept for staging such a transformative research strategy, a highly sensitive quantitative *in vivo* imaging methodology targeting human CD8^+^ cells is described, with a particular interest towards studying the immunobiology of CD8^+^ T cells as the major population of human CD8^+^ cells. For this, the recently developed imaging probe ^89^Zr-Df-Crefmirlimab also known as ^89^Zr-Df-IAB22M2C, is used with positron emission tomography (PET). IAB22M2C is a biologically inert 80-kDa minibody with high affinity to human CD8 and has accelerated serum clearance compared to full-sized antibodies, making it particularly favorable for *in vivo* imaging. IAB22M2C conjugated to the chelator desferrioxamine (Df) and radiolabeled with Zirconium-89 (^89^Zr) has been successfully used in a number of preclinical and clinical trials with a focus on cancer immunotherapy applications *(26,27)*. With a long radioactive half-life of 78.4 hours, ^89^Zr allows the tracer’s biodistribution to be followed for several days post injection (p.i.). However, because of its long half-life, radiation dose concerns have prevented wider application of ^89^Zr-immunoPET in non-life-threatening disease and healthy populations. The advent of total-body PET scanners covering all or much of the body *(28)*, which offer a radiation detection sensitivity increase of 1‒2 orders of magnitude compared to conventional PET scanners *(29)*, enables high signal-to-noise ratio imaging of ^89^Zr-based radiotracers at substantially lower injected doses in addition to capturing kinetics across all the organs and tissues of interest. This enables complete characterization of the pharmacokinetics of these immunological PET tracers across a wide range of applications *(28,30)*. Total-body PET is currently the only available technology that allows noninvasive *in vivo* measurements of T cell distribution and kinetics inside all tissues in human subjects, with acceptable radiation dose burden.

In this work, low (< 20 MBq) doses of ^89^Zr-Df-Crefmirlimab tracer were used with the 194-cm-long uEXPLORER total-body PET scanner to study the biodistribution and kinetics of CD8^+^ cells in COVID-19 convalescent patients and in healthy controls.

## RESULTS

Eight subjects were enrolled, including five COVID-19 convalescent patients and three healthy controls. The demographics of the study participants are included in Table 1. All subjects had received at least one dose of a COVID-19 mRNA vaccine prior to their first imaging session, except for one healthy subject (Sub08) who had not received any vaccination prior or during the imaging study. The participants varied in vaccination timeline with respect to their imaging sessions and one patient (Sub01) contracted COVID-19 prior to vaccination. COVID-19 patients were possibly exposed to different variants of the virus with infection timelines ranging from January 2021 to March 2022 and varied in terms of infection symptoms and past medical history (Table S1). The last two enrolled patients (Sub04 and Sub05) had the mildest symptoms, with exposure timelines during or after the Omicron variant surge in California.

**Table 1.** Demographics of study participants.

| Demographic | COVID-19 | Control |
| --- | --- | --- |
| Total participants (n) | 5 | 3 |
| Age range (y) | 27–51 | 25–59 |
| Sex (n) |  |  |
| Male | 0 | 2 |
| Female | 5 | 1 |
| BMI range ($\text{kg}/\text{m}^2$) | 20–43 | 21–31 |

The mean injected radiotracer activity was 18.8 MBq (0.51 mCi), with a range of 15.4–21.8 MBq (0.42–0.59 mCi). The mean injected minibody mass was 1.50 mg, with a range of 1.33–1.77 mg. The injections and PET imaging were well tolerated, with no adverse reactions to the infusion. No adverse effects and no clinically significant changes in vital signs were observed during the study. Two COVID-19 patients did not have dynamic scans and the dynamic scan of one control subject was terminated early at 65 min due to patient motion and discomfort.

### Blood Clearance of the Radiotracer

The whole-blood clearance was best described by a triexponential model, with an initial half-life of 5.1±2.2 min (range, 2.6–8.1 min), an intermediate half-life of 55.9±28.0 min (range, 29.3– 120.0), and a terminal elimination half-life of 22.1±11.8 h (range, 13.4–50.4 h). One COVID-19 subject (Sub02) showed significantly longer terminal half-life compared to all other subjects (Fig. S1).

### Total-Body Biodistribution of the Radiotracer

Standardized uptake value (SUV) images of the baseline scans of COVID-19 convalescent patients and healthy controls at three imaging timepoints showed high uptake in lymphoid organs of all subjects (Fig. 1), with the highest uptake observed in the spleen of all subjects, followed by bone marrow, liver, tonsils, and lymph nodes. Bone marrow uptake was particularly prominent in the vertebrae, sacrum, ilium, ribs, sternum, clavicle, and scapulae of all subjects and showed variable extended lengths in humeral and femoral shafts. Peripheral lymph nodes showed marked uptake in all subjects as early as 30‒90-min p.i. and peaked at the 48-h timepoint. Prominent uptake was observed in head and neck lymph nodes of all subjects and a subset of subjects also showed uptake in their axillary, pelvic, mediastinal, as well as upper and lower limb lymph nodes. Consistent with the expected hepatobiliary clearance of the radiotracer, gallbladder was visualized during the dynamic scans of all subjects, except for the two COVID-19 subjects who did not have dynamic scans, one of which (Sub03) had a history of cholecystectomy. Excluding Sub03, gallbladder was still visualized in 9 out of 11 scans performed at each of the 6-h and 48-h timepoints. Furthermore, subjects showed activity in their bowel during the 48-h time-course of the study, with the largest activity observed in the colon and rectum. Cross sectional analysis suggested the activity was in the large bowel lumen, supported by changes in the location of the activity over the 48 h. The small bowel also contained low levels of activity; however, it was not possible to confirm whether the activity was in the lumen or in the Peyer’s patches of the small bowel wall. Muscle, cerebrum, and cerebellum uptakes were low in all subjects, with SUVmean values below 0.2, 0.7, and 0.8, respectively, at all timepoints. Lymphoid tissues in the nasal and pharyngeal area had visible uptake in all subjects with similar range of values. Comparing the baseline images of the COVID-19 patients to the images from their 4-month follow-up scans (Fig. 2) showed that despite the observed differences and heterogeneity among the study participants, irrespective of their study group, the follow-up scans of each patient exhibit striking similarities to their baseline scans particularly in bone marrow.

**Fig. 1.**
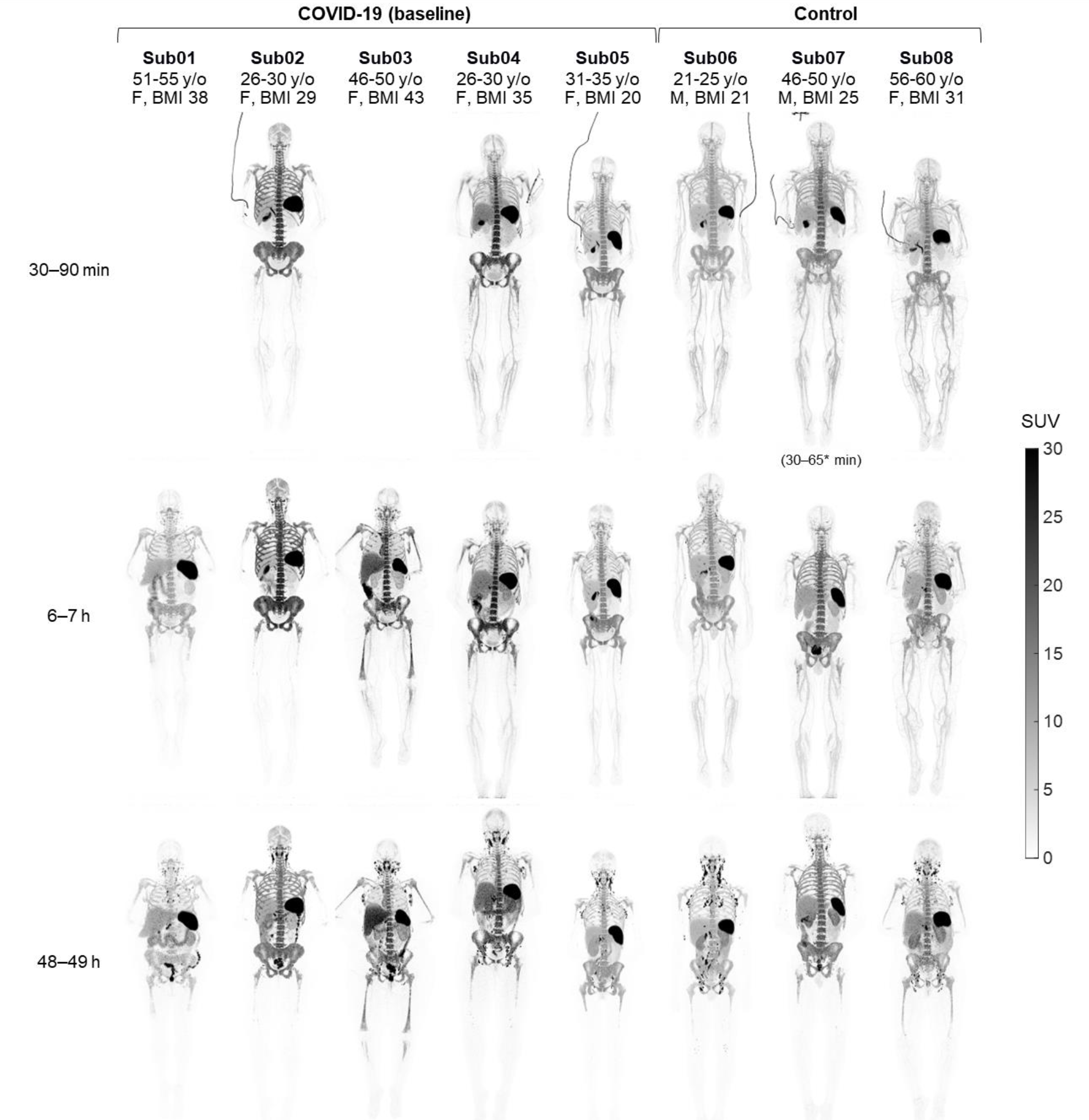
Maximum intensity projection (MIP) of decay-corrected SUV images of the baseline scans. The baseline scans of COVID-19 convalescent patients and healthy control subjects are compared at three imaging timepoints. Sub01 and Sub03 skipped dynamic imaging.

**Fig. 2.**
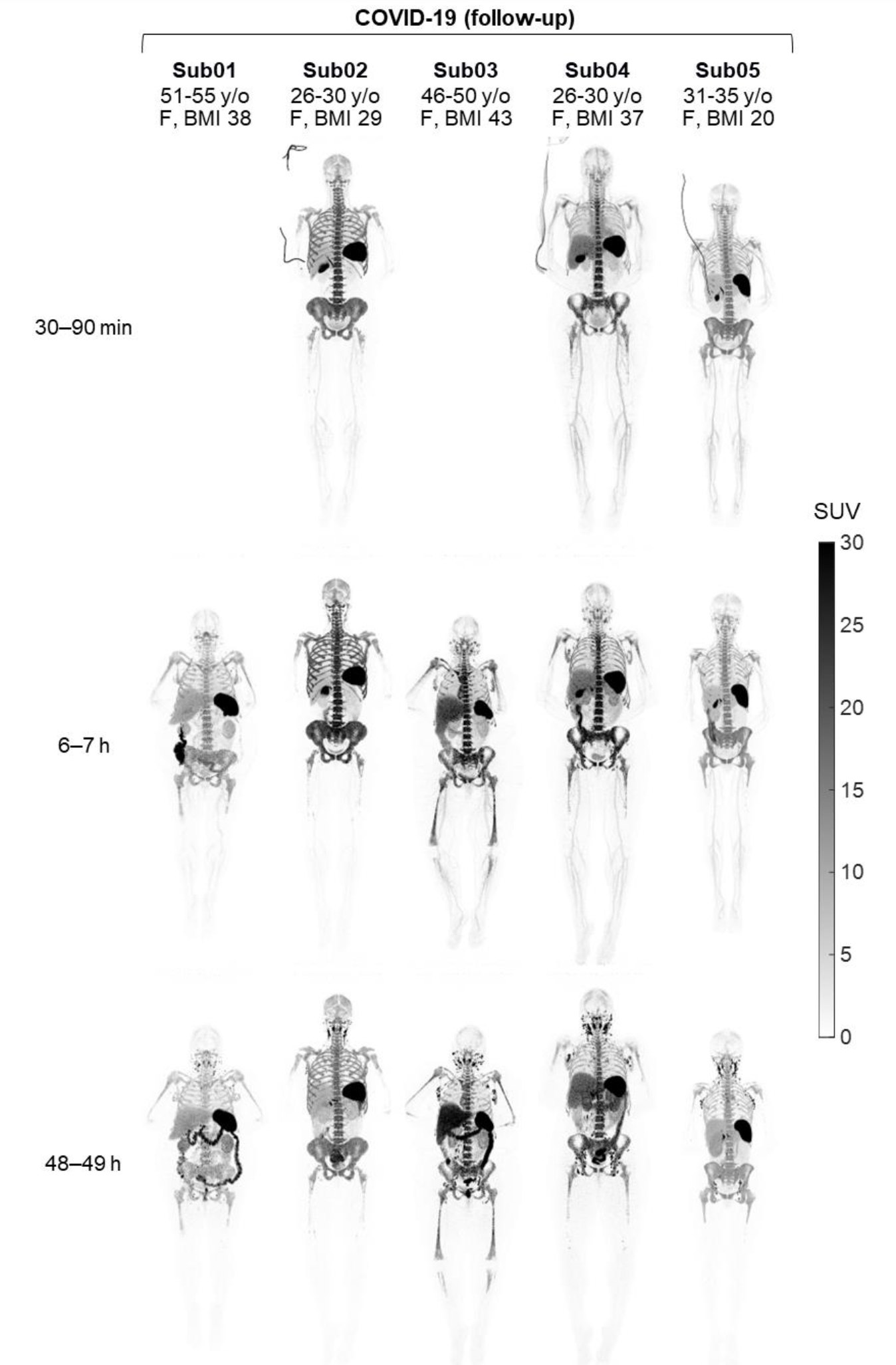
MIP of decay-corrected SUV images of the 4-month follow-up scans. The follow-up scans of the COVID-19 convalescent patients are shown at three imaging timepoints. Sub01 and Sub03 skipped dynamic imaging.

### Kinetics During the 48-h of Imaging

The time activity curves (TACs) from all investigated organs of interest showed consistent trends in tracer kinetics in all subjects (Fig. 3). Different regions of bone marrow showed an increasing trend during the 90-min of the dynamic scans in all subjects, with a plateauing rate of uptake towards 90 min. Spleen TACs also showed a plateauing increasing trend during the 90-min of the dynamic scans, except in two COVID-19 subjects (Sub02 and Sub04), in which the TACs showed a decrease after peaking at around ∼1-h. Lymph nodes visualized with high contrast at 6-h and 48-h timepoints, could mostly be visualized also on the 30-90 min images with SUVpeak values in the range of 0.2‒5.2. Between the 6-h to 48-h timepoints, all subjects showed a decrease of SUV in spleen, bone marrow, and lungs and parallel increase of SUV in lymph nodes and tonsils, which was quantified by percentage change in SUV at the 48-h timepoint relative to the 6-h timepoint in all investigated organs-of-interest (Fig. S3). Comparing the percentage change in SUV during the last 42 h showed similar trends in all organs-of-interest in all subjects, with no significant difference between the COVID-19 and the control group. Liver uptake showed relatively smaller changes during the last 42-h, with an increasing trend in three subjects.

**Fig. 3.**
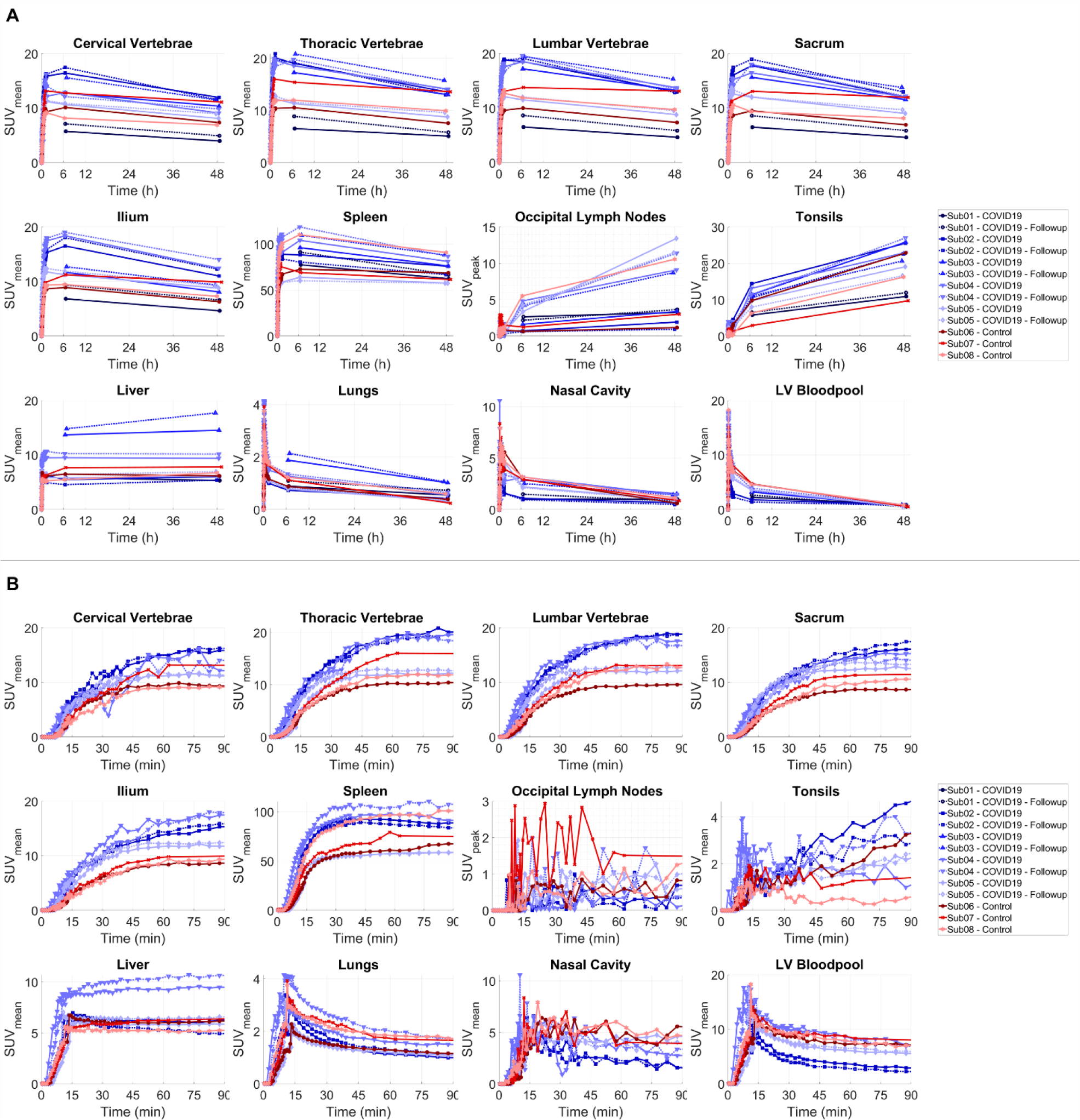
Decay-corrected TACs of different organs of interest. (**A**) The TACs representing the delivery, retention, and clearance of the tracer over the 48-h time course of the study are shown for bone marrow (cervical, thoracic, and lumbar vertebrae, sacrum, and ilium), spleen, liver, lymph nodes, tonsils, lungs, nasal cavity, and the left ventricle (LV) blood pool, in addition to (**B**) zoomed-in plots on the first 90-mins after tracer administration for all subjects. Control and COVID-19 subjects are in shades of red and blue, respectively. The lymph node TACs show an example lymph node selected from the occipital region of each subject (PET/CT images shown in Fig. S2). The occipital region was selected as a common area where all subjects showed quantifiable uptake and the TACs were not affected by spill-over from adjacent lymph nodes or blood vessels.

Tissue-to-blood ratio (TBR) curves plotted as a function of time for different regions of bone marrow and liver showed a separation between the COVID-19 and the control group during the 90-min dynamic scans up to the 7-h timepoint, with higher values observed in the COVID-19 group (Fig. S4). Sacrum and ilium bone marrow showed the largest differences between the two groups. Comparing the TBRs of all subjects at 30‒90 min and 6‒7 h timepoints (Fig. 4) showed significant differences between the COVID-19 and the control group at the 6‒7 h timepoint in liver, different bone marrow regions, and tonsils (p=0.036). Moreover, TBRs of all bone marrow regions, spleen, and tonsils were 2‒3 times higher in one COVID-19 subject (Sub02) than all other patients during the first 7 h. No significant difference was observed in spleen, lungs, or nasal cavity between the two groups.

**Fig. 4.**
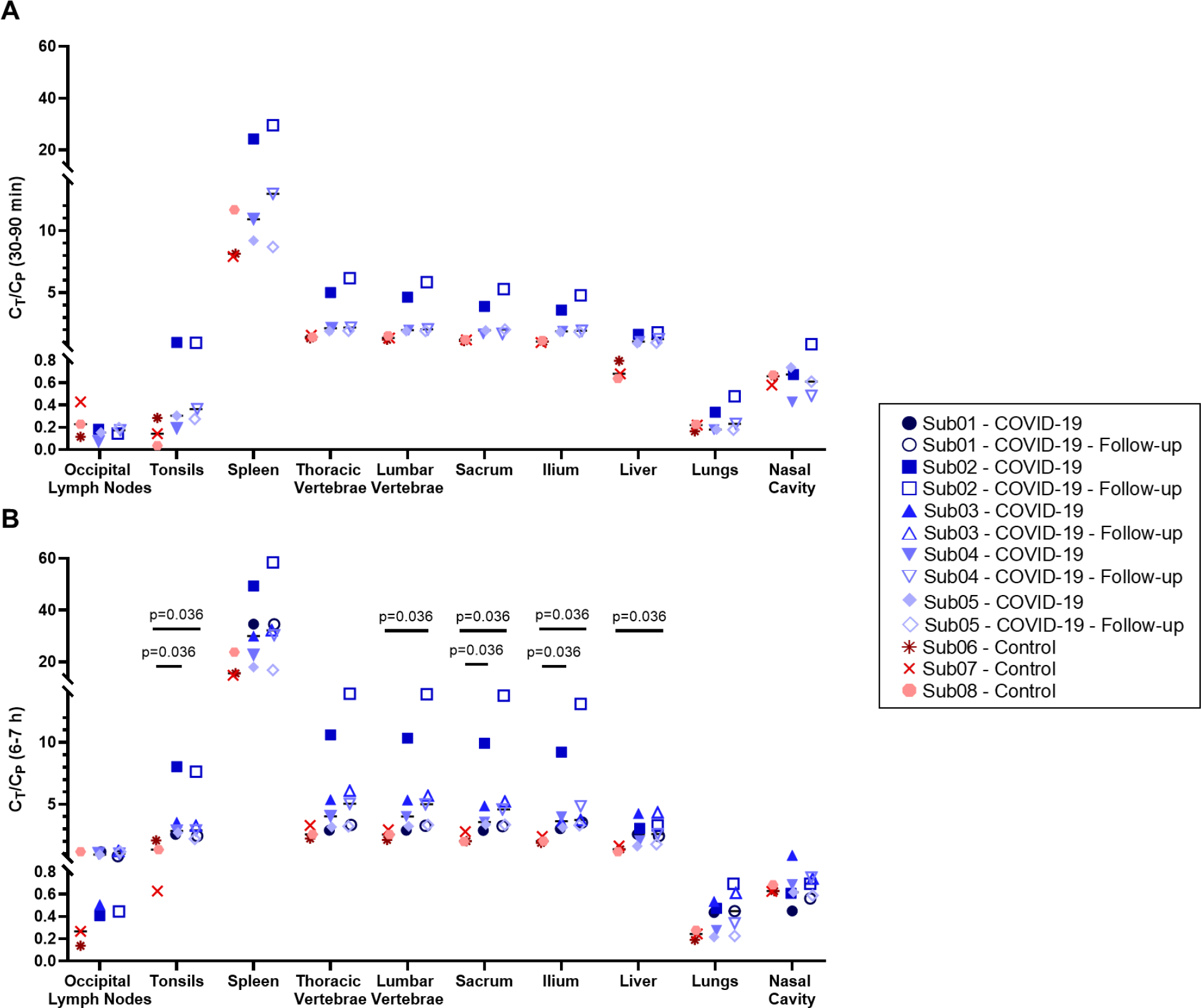
TBRs of different organs-of-interest. TBRs are compared between COVID-19 and control subjects in lymph nodes, tonsils, spleen, bone marrow, liver, lungs, and nasal cavity (**A**) during the 30‒90 min of the dynamic scans and (**B**) at the 6‒7 h timepoint.

Comparing the percentage changes of TBR at 4-month follow-up scans relative to the baseline scans of the COVID-19 patients (Fig. 5) did not show consistent trends in most organs-of-interest, except for the bone marrow, in which a consistent trend towards increased TBR was observed in all bone marrow regions of the COVID-19 subjects during the first 7 h of imaging. While this increasing trend in bone marrow TBRs ranged from 2‒42% at the 6-h timepoint in 4 out of 5 COVID-19 subjects, in one COVID-19 subject (Sub05), the follow-up scans did not show substantial changes in the bone marrow regions, with only 0‒5% changes compared to the baseline scan.

**Fig. 5.**
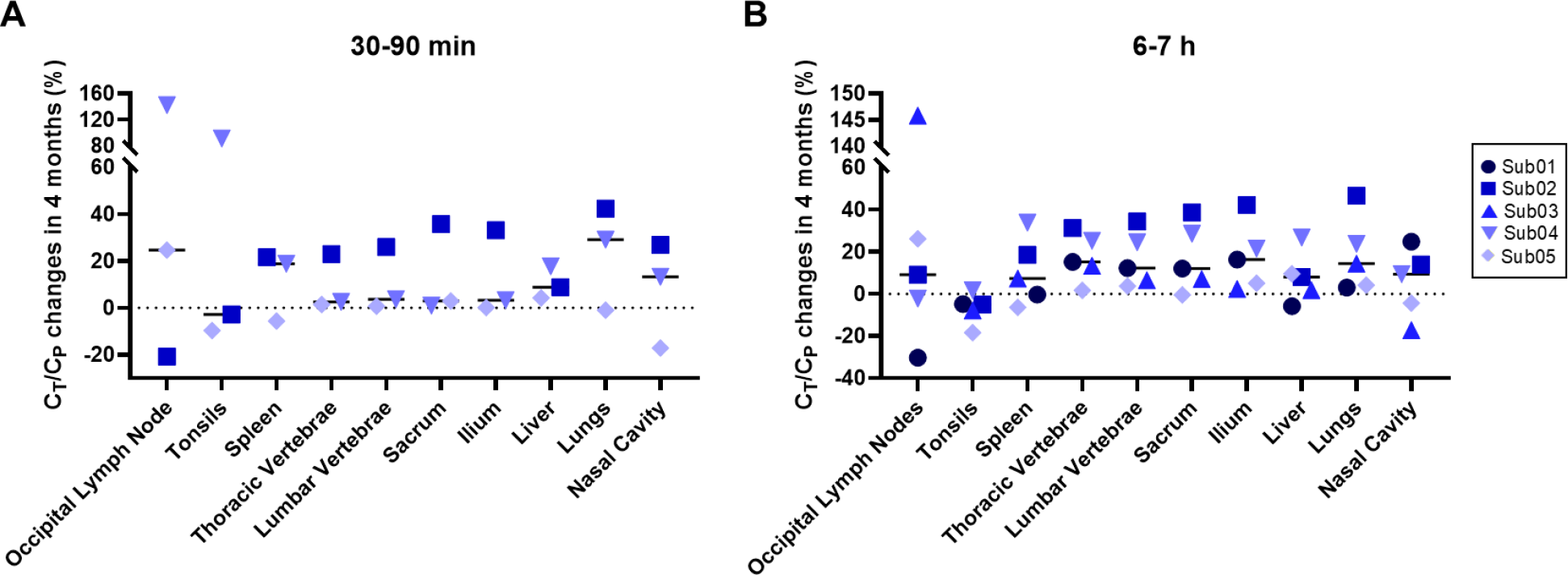
Longitudinal changes of TBRs in different organs-of-interest of COVID-19 convalescent patients. Percentage changes of TBR at 4-month follow-up scans of the COVID-19 patients relative to their baseline scans is shown in lymph nodes, tonsils, spleen, bone marrow, liver, lungs, and nasal cavity at (**A**) 30‒90 min, and (**B**) 6‒7 h.

### Kinetic Modeling Approaches for ^89^Zr-Df-Crefmirlimab

Conventional PET kinetic modeling approaches based on one-tissue and two-tissue compartmental models with Akaike information criterion (AIC) model selection successfully fitted the TACs in lungs, spleen, bone marrow, tonsils, and selected occipital lymph nodes (Fig. S5 and Table S2). In all cases, all available timepoints were used for model fitting and using the earlier timepoints alone was not sufficient to accurately model the TACs at later timepoints. Visual inspection of the AIC-preferred model fits in all organs-of-interest suggested low residual errors in fitting all imaging timepoints (Fig. S6 and Fig. S7), except for the lymph nodes, in which TACs were more affected by statistical noise due to small number of voxels included in the analysis. Normalized sensitivity plots (Fig. S8) showed increasing sensitivity of all model microparameters up to the 48-h timepoint, except for *v*_*b*_ and *K*_1_, which reached their maximum sensitivity at earlier timepoints in some cases. Correlations observed between microparameters of the 2T model (Table S3) were mostly expected and similar to those commonly observed in conventional PET kinetic modeling. Simulations of the TAC noise model in all organs-of-interest showed low biases in all microparameter estimations (Table S4), suggesting small effects from statistical noise and high confidence in microparameter estimation at organ-level; with the exception of lungs that showed increased errors compared to other organs. In all investigated organs, the slope of the Patlak plot changed from 90-min to 48-h and since only two datapoints were available at later timepoints, it was not possible to determine the equilibrium time (Fig. S9).

In lungs, AIC favored the 2T model over 1T3P model in all subjects, however, the AIC values were close between 2T4P and 2T5P models in many cases (Fig. S5). All three models could fit the early 90-min data points, but the fitting errors were larger for the late timepoint data with the 1T3P model (Fig. S6 and Fig. S7). Air fraction correction was not applied to the results, as using the low-dose CT images resulted in overestimation of air fraction values, particularly in high-BMI subjects. With no air fraction correction, no significant difference was observed between the control and COVID-19 groups and only one COVID-19 patient (Sub02) showed increased *K*_*i*_ values.

In spleen, AIC largely favored the 2T5P model in all subjects (Fig. S5). 2T4P and 1T3P models could not fit the later timepoints and increasing the weighting factors of the late timepoints resulted in inaccurate fits on the first 90-min data (Fig. S10). *K*_1_ and *k*_2_ were highly correlated (>98%) with *v*_*b*_ and increase in *K*_1_ up to its upper bound was compensated by a decrease in *v*_*b*_. Therefore, *v*_*b*_ was set to 0.4 during the fitting in all cases. The fitted model suggested that the concentration of the bound tracer in the second compartment increases up to 12‒24 h p.i. and starts to drop thereafter (Fig. S11). No significant difference was observed between the two groups.

In bone marrow, AIC favored the 2T5P model in all subjects (Fig. S5). The 2T4P model could fit the data from all timepoints, but with higher AIC values, whereas the 1T3P model could not fit the 48-h timepoint. The microparameter estimates for the 2T5P model were similar in sacrum and ilium. 2T5P model *K*_*i*_ values were higher in bone marrow of COVID-19 patients compared to the controls (p=0.1) (Fig. 6). Similar to spleen, the fitted 2T5P model suggested increasing concentration of the bound tracer up to 12‒24 h p.i. and a decrease thereafter.

**Fig. 6.**
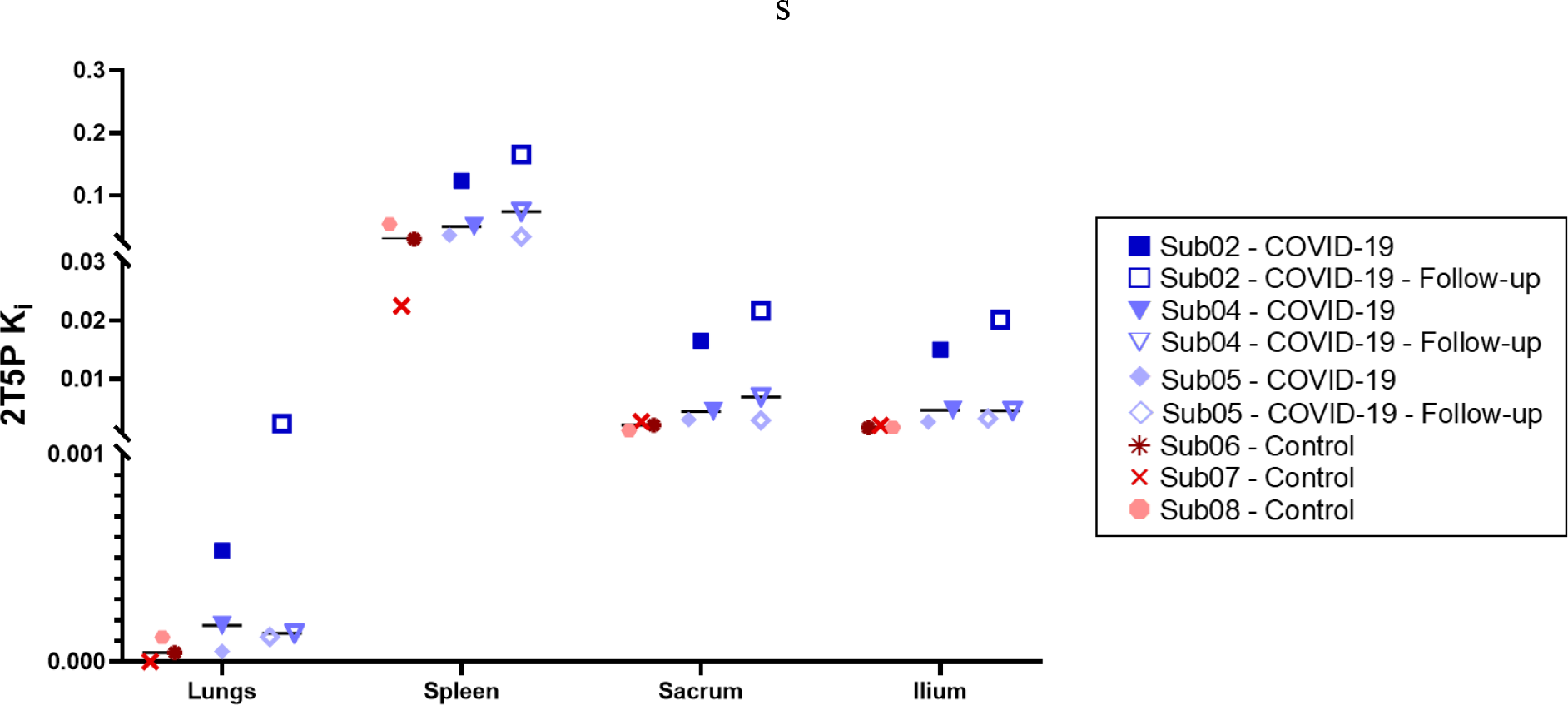
Net influx rate of the 2T5P model. Net influx rate (***K***_***i***_) obtained by 2T5P model fits on lungs, spleen, and sacrum and ilium bone marrow are compared in all subjects with dynamic scans.

In tonsils, AIC favored the 1T3P model and in lymph nodes, AIC favored the 2T4P and 1T3P models interchangeably (Fig. S5). In many cases, the 2T model microparameters were zero or resulted in inconsistent estimates, therefore, 1T3P model was selected for the lymph nodes as well.

### Uptake in Thymus

Thymus uptake was observed only at the 48-h timepoint in two subjects (Fig. 7), including one COVID-19 convalescent patient (in both baseline and follow-up scans) and one control subject, with SUVmean values of 4.0 and 4.7, respectively. The COVID-19 and control subject were both below 35 years old and had the lowest BMIs among all subjects, with BMIs of 20 and 21 kg/m^2^, respectively. Thymus fatty degeneration scoring based on low-dose CT images in all subjects (Fig. S12) showed solid thymic gland with Score 3 in the COVID-19 subject and half fatty and half soft-tissue attenuation with Score 2 in the control subjects. Two other under-30 y/o subjects in the study (Sub02 and Sub04) also showed solid thymic glands with Score 3. Both subjects were in the COVID-19 group and had higher BMIs compared the two subjects that showed thymus uptake in their PET images.

**Fig. 7.**
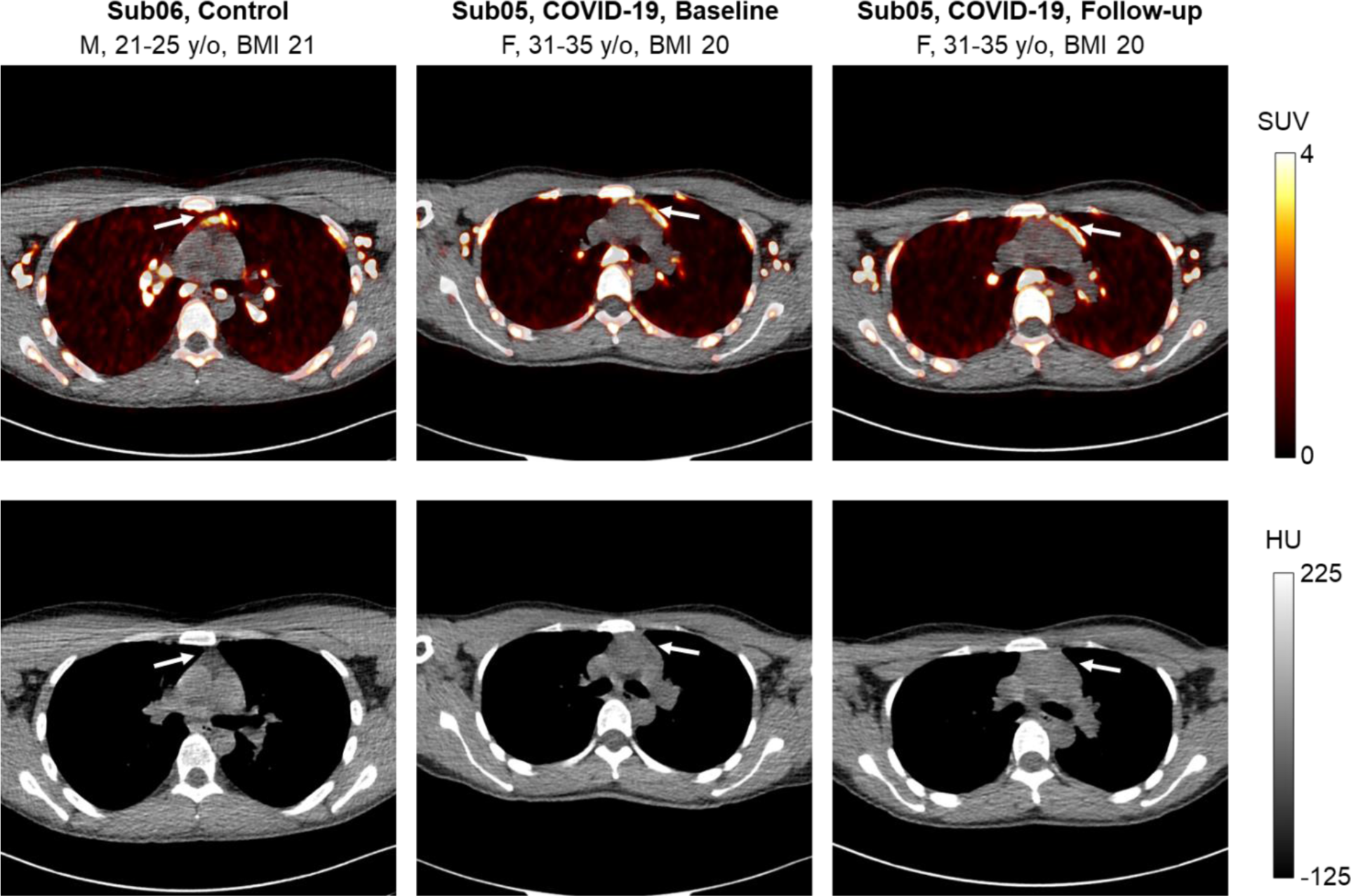
PET/CT image slices of thymus uptake. Selected transverse PET/CT image slices of one control subject and one COVID-19 patient (at baseline and follow-up scan) showing thymus uptake at 48-h timepoint of imaging.

### Peripheral Blood Assays

CD8^+^ and CD4 T^+^ cell immunophenotyping results (Fig. 8 and Fig. S13, respectively) showed an increase in percentage of CD8^+^ T cells and a decrease in percentage of CD4^+^ T cells in COVID-19 convalescent patients compared to the controls (p=0.036). Moreover, an increased frequency of activated CD8^+^ T cells was observed in COVID-19 convalescent patients, both in CD38^+^HLA-DR^+^ cells and CD56^+^ cells (p=0.036). No difference was observed in frequency of PD-1^+^ cells between the two groups.

**Fig. 8.**
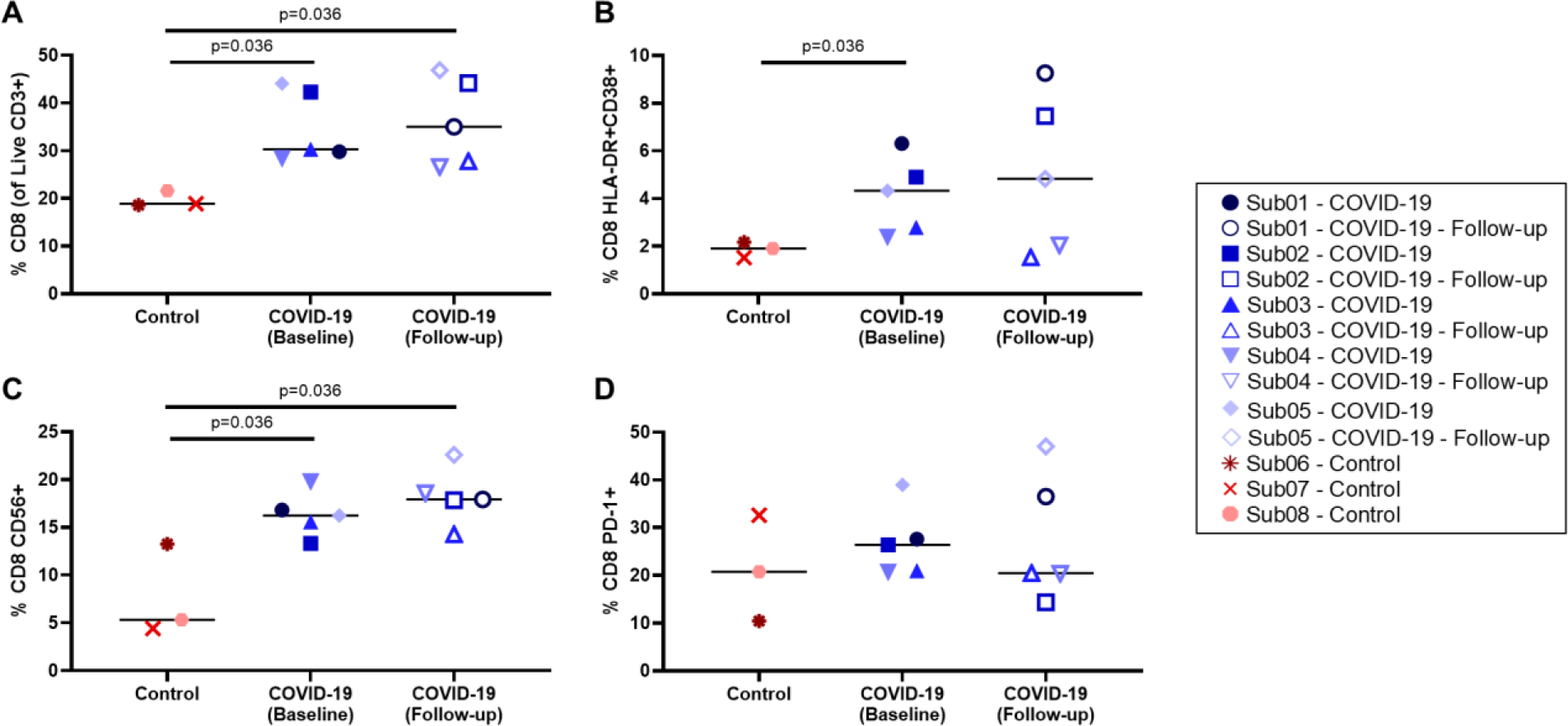
Peripheral blood CD8^+^ T cell phenotyping. (**A**) Percentage of CD8^**+**^ T cells within the live CD3^+^ population, (**B**) percentage of activated CD8^**+**^ T cells characterized by CD38 and HLA-DR co-expression and (**C**) CD56 expression, and (**D**) percentage of exhausted CD8^**+**^ T cells characterized by PD-1 expression are compared in all subjects.

Comparing the percentage of memory subsets of CD8^+^ and CD4^+^ T cells in all subjects (Fig. S14). showed a trend towards higher frequency of CD8^+^ effector memory T cells in the COVID-19 convalescent individuals compared to the controls. Percentage of mucosa-associated invariant T (MAIT) cells, regulatory T (Treg) cells, natural killer (NK) cells, and B cells compared between the two groups (Fig. S15) showed a trend towards higher percentage of MAIT cells in COVID-19 convalescent patients (p=0.072) and no significant difference was observed between the two groups with other cell types.

Total percentage of CD8^+^ and CD4^+^ memory T cells responding in any way (CD107a, IFNγ, IL2, MIP-1β, or TNFα) to SARS-CoV-2 spike and nucleocapsid proteins compared in all subjects (Fig. S16) showed trends toward higher magnitude CD8^+^ and CD4^+^ responses to both spike and nucleocapsid in COVID-19 convalescent participants compared to the controls, which was significant for the spike-specific CD4^+^ responses (p=0.036), but no significant differences were observed between the baseline and 4-month follow up scans. Furthermore, CD4^+^ responses were of higher magnitude than CD8^+^ responses. Individual responses in CD8^+^ and CD4^+^ memory T cells (Fig. S17 and Fig. S18, respectively) showed potential trends towards increasing spike-specific CD8^+^ T cell degranulation response (CD107a^+^), higher frequencies of spike-specific CD4^+^ IL2^+^ T cells, and higher frequencies of spike-specific CD4^+^ IFNγ^+^ T cells in COVID-19 convalescent patients (p=0.036). Lastly, SARS-CoV-2 specific CD8^+^ and CD4^+^ responses were mainly dual and monofunctional (Fig. S19 and Fig. S20, respectively) and CD4^+^ T cell responses were slightly more polyfunctional than CD8^+^ responses. Only a slight increase in polyfunctionality could be observed for nucleocapsid-specific CD8^+^ T cells in COVID-19 convalescent participants at the 4-month timepoint.

## DISCUSSION

We present the first results on the biodistribution and kinetics of a CD8-targeted radiotracer in healthy control subjects and in subjects following a viral infection. The biodistribution and general trends were in good agreement with previous oncologic human studies using ^89^Zr-Df-Crefmirlimab *(26,27)*. Although there was a 6-fold reduction in injected radiation dose compared to previous Phase I human studies, significant image quality improvements, both in terms of noise reduction and higher contrast in small structures, were observed in the total-body PET scans compared to the previous human PET scans *(26,27)*. Notably, a large number of high-contrast lymph nodes were visualized in all subjects and the tracer kinetics in any given organ followed consistent trends in all subjects. Considering the low positron fraction of ^89^Zr, the remarkable image quality obtained in this study demonstrated feasibility of high-quality dynamic imaging with ^89^Zr-labeled immunological tracers across the whole body, at doses that permit longitudinal imaging in healthy subjects and any disease state.

This study revealed significant differences in the bone marrow CD8 concentrations of COVID-19 convalescent patients compared to controls, with increased TBRs in the first 7-h of the study and increased net influx rates (2T5P *K*_*i*_) observed in the bone marrow of COVID-19 patients. Such changes were not evident in the commonly used SUV images, because the measured SUV cannot account for the time-varying tracer concentration in the blood and tissue compartments, nor the effects of cell trafficking. This study demonstrates the role of dynamic imaging and kinetic modelling in providing quantitative biomarkers. While SUV TACs from the first 90 min showed higher values in sacrum and ilium bone marrow of the COVID-19 patients compared to controls, SUV TACs overlapped between the two groups for other bone marrow regions, particularly at later timepoints where the two additional COVID-19 subjects with no dynamic scans were included. Plotting TBRs as a function of time however showed more distinct differences between the two groups in all bone marrow regions up to the 7-h timepoint. The tissue-to-blood ratio curves from the three control subjects showed very similar values particularly in sacrum and ilium; while within the COVID-19 group, one subject (Sub02), who got infected with COVID-19 twice, showed consistently significant higher values in all bone marrow regions compared to all subjects. Other COVID-19 patients showed a separate cluster of results, but still higher than the controls. Tumor-to-blood ratio has been previously shown to be a surrogate for the net metabolic rate of other radiotracers such as ^18^F-fluorodeoxyglucose in tumors *(31)*. Comparing TBRs among different subjects at a specific timepoint, while equilibrium has not been reached, requires extra care, as TBR changes as a function of time and is affected by blood clearance rates. Plotting TBR as a function of normalized time in Patlak plots accounts for variations in blood clearance. The similarity of the TBR plots (Fig. 3) to Patlak plots (Fig. S9) during the first 7 h is consistent with similar initial and intermediate blood clearance rates (Fig. S1) among all subjects. Patlak plots also reflect the differences in terminal blood clearances and show improved separation between the two groups at later timepoints. Furthermore, TBRs from the 6-h timepoint were highly correlated with net influx rates (*K*_*i*_) obtained from 2T5P fits on the complete 48-h TACs (ρ>0.99 in bone marrow and spleen and ρ>0.93 in lungs), suggesting that they can be used as surrogates for *K*_*i*_. The mitigated separation between the two groups at later timepoints in the Patlak plots may be attributed to increased cell trafficking effects that are not accounted for. Future studies should investigate correlation of early-timepoint TBRs with model macro-parameters, best describing the CD8 tissue density in absence of cell trafficking effects.

In addition to higher CD8 TBRs values observed in bone marrow of COVID-19 convalescent patients compared to controls, flow cytometry results also showed higher percentage of CD8^+^ T cells, higher percentage of activated CD8^+^ T cells, and higher percentage of CD8^+^ memory T cells in peripheral blood of COVID-19 convalescent patients compared to controls. Furthermore, the two longitudinal scans performed on COVID-19 patients showed consistently increasing trends in the bone marrow of all subjects between 2- and 6-months post-infection. While the magnitude of these longitudinal changes varied among different patients, low degree of heterogeneity was observed in different bone marrow regions of each patient. The peripheral blood flow cytometry data on the other hand only showed a non-significant population-based increase in the median of the 4-month follow-up scans compared to the baseline scans. Although a high degree of heterogeneity can be expected among human subjects, the large variations observed within each group may also be partly due to the limited precision of the flow cytometry methods, which usually require larger cohort of subjects for making statistically meaningful conclusions. Such limitations suggests that flow cytometry methods may be inadequate for studying small longitudinal changes in individual subjects. These large variations can be also observed in the reported results from previous studies using peripheral blood of COVID-19 patients, in which a general trend towards declining SARS-CoV-2 specific CD8^+^ memory T cell response is observed from statistical analysis on peripheral blood of two large cohort of patients between 1‒8 months post infection, but inconsistent findings are observed in longitudinal measurements in individual subjects *(9,10)*. The small number of participants in this pilot study makes it difficult to conclude whether the observed trends in increased percentages of CD8^+^ T cells in the peripheral blood of COVID-19 patients at the 4-month follow up scans are due to the heterogeneity of COVID-19 patients, or clinical differences among the subjects of this study with those of previous studies, or due to methodological variations. Furthermore, while vaccination timeline is expected to affect the spike-specific responses, some inconsistencies can be observed in SARS-CoV-2 specific responses, such as the high spike-specific and nucleocapsid-specific responses in the unvaccinated control subject (Sub08), which could be due to cross-reactivity with seasonal Coronaviruses or possible subclinical infection or exposure. Nevertheless, image-based results show no significant difference in bone marrow of the unvaccinated subject compared to other control subjects, but higher TBRs, in similar range to COVID-19 subjects, can be observed in the spleen of the unvaccinated subject, which needs further investigation.

Kinetic modeling based on conventional PET compartmental models showed reproducible results at organ-level, which were in good agreement with immunobiology of the investigated organs and the expected T cell trafficking effects were reflected in the kinetic model selection results. Although the conventionally used one-tissue and two-tissue compartmental models do not include separate pathways for the trafficking of radiolabeled cells and therefore, they are not expected to accurately represent the kinetics of the ^89^Zr-Df-Crefmirlimab in lymphoid tissue, in the absence of cell trafficking, the main mechanism of uptake can be simplified and approximated by the 2T4P model, in which upon entrance of the free ^89^Zr-labelled minibody tracer into tissue, the tracer binds to the CD8 receptors and gets irreversibly internalized within the cell. In the presence of cell trafficking, which is non-negligible over the 0‒48-h timeframe in lymphoid organs, all rate constants of tracer exchange between the model compartments will include components from trafficking and the model will require a non-zero *k*_4_ to represent the trafficking of radiolabeled CD8^+^ T cells out of the tissue. This is reflected in the AIC model selection results for spleen, bone marrow, and lungs. The 2T5P model, which includes a non-zero *k*_4_, is largely favored in spleen, where a large population of naïve recirculating CD8^+^ T cells are expected and the migration of radiolabeled CD8^+^ T cells out of spleen may prominently contribute to the *k*_4_. A similar effect can be observed in bone marrow, but to a lesser extent. In the case of healthy lung tissue, relatively low concentrations of CD8^+^ T cells are expected and a large fraction of the CD8^+^ T cell population could be non-circulating tissue-resident T cells. As a result, the difference between the 2T5P and 2T4P models becomes much smaller, particularly in the healthy subjects where 2T4P is favored, which could be attributed to a smaller effect from T cell trafficking in such cases.

Furthermore, the effects of blood flow and permeability of the blood vessels to the 80-kDa minibody molecules in different types of tissue could explain the differences observed in the TACs of different organs and their corresponding tissue compartments (Fig. S11). The presence of sinusoidal capillaries in spleen and bone marrow, which allow exchange of large molecules between blood and the surrounding tissue, in addition to the high blood flow in these two CD8-rich organs results in fast entrance of the free tracer into the tissue followed by a slower process of binding within the tissue in the second tissue compartment. While a continuous exchange of labeled CD8^+^ cells between tissue and blood is present in parallel, the high permeability and blood flow could result in saturation of binding sites in these two organs as reported in a previous dose escalation study when a 1.5 mg minibody mass dose is used *(27)*. The following decrease in the concentration of the second tissue compartment (Fig. S11) can therefore be attributed to the flux of radiolabeled CD8^+^ T cells out of the tissue. In the case of lungs, lower permeability, lower blood flow, and lower concentrations of CD8^+^ T cells can be expected than spleen or bone marrow, and as a result, a large fraction of the initial signal in the lungs is due to tissue blood fraction and low levels of free tracer entering the tissue, followed by slow binding and clearance. In the case of lymph nodes and tonsils, where high concentrations of binding sites are expected but tracer delivery to the tissue may be slower due to significantly lower blood flow compared to spleen, bone marrow, and lungs, AIC favors the 1T3P model in which the low amounts of free tracer slowly entering the tissue bind relatively quickly to the highly available binding sites in the tissue. However, the increase of uptake in lymph nodes and tonsils over time may also be largely attributed to migration of radiolabeled T cells to these tissues, which cannot be separated from the free tracer uptake in the current model. It should be noted that the whole-blood compartment itself contains CD8^+^ cells and as these cells get radiolabeled with the tracer and are in continuous exchange with the radiolabeled CD8^+^ cells in different lymphoid organs, the concentration of radiolabeled CD8^+^ cells in whole-blood changes as a function of time. Future studies should investigate these changes in the whole-blood compartment as a function of time and incorporate them either directly in the compartmental model or indirectly with correction factors, in order to separate the free tracer uptake kinetics from cell trafficking kinetics. In the case of lymph nodes in particular, a second distinct uptake pathway may also be present through afferent lymphatic vessels, which needs further detailed investigation. Previous studies on the anatomy of the lymphoid compartment within lymph nodes have described a 70-kDa size-exclusion limit restricting access of heavy-weight-molecules to lymph node T cell zones through lymph *(32)*. However, there has been ongoing research on identifying special routes of transport for high-molecular-weight molecules into the conduit system of the lymph nodes *(33)*.

Normalized sensitivity plots suggest that 48 h of imaging is required for estimating the model microparameters and might be sufficient. However, more intermediate imaging timepoints are required to study this in detail. The Patlak plots further illustrate this need, where even in the case of the healthy lung tissue in which AIC favors the 2T4P model, the equilibrium time seems to be larger than 7-h and cannot be determined from the current dataset. Furthermore, although simulations of the TAC noise model show very promising results, particularly in spleen and bone marrow with low bias values in the range of ±1% for most microparameters, it is still possible that the noise model was underestimated for the later timepoints, since only two datapoints were available for fitting and the model fits might have been affected by overfitting. This can also be investigated in future studies with more intermediate imaging timepoints.

While the kinetics of the radiotracer are substantially different in the liver compared to bone marrow and conventionally, liver is not considered a lymphoid organ, TBR curves in the liver also show higher values in the COVID-19 subjects compared to the controls and substantially higher values for Sub02. This could be due to the presence of subsets of CD8^+^ T cells in the liver, the population of which may be affected during viral infection. However, due to the hepatobiliary clearance of the radiotracer and dual blood supply of liver, further studies are required to accurately model the kinetics of the tracer in the liver and quantify the cell trafficking effects. Furthermore, TBR curves in the lungs show overlapping curves between the two groups, which may be expected since as all COVID-19 patients in the study had relatively mild infection, with no hospitalization and no findings on lung involvement. However, no certain conclusions can be made as air fraction correction was not applied to the data due to low-quality of CT acquisitions. The systematic significant differences observed in all tissue concentrations of one COVID-19 patient (Sub02), which is not evident in the peripheral blood results, need further investigation with respect to their clinical record and history of two COVID-19 infections, including an asymptomatic infection after one vaccination dose and a second infection after 3 months with significant illness and respiratory symptoms.

The thymus uptake observed in the two lowest-BMI subjects, both under 30 y/o, is consistent with previous reports on increased fatty degeneration in thymus with aging and high BMI *(34)*. While fatty degeneration scoring based on CT images shows predominant soft-tissue attenuation in two higher-BMI subjects under 30 y/o, with Score 3, no thymus uptake is observed in their corresponding PET images. Furthermore, observing the thymus uptake only at the 48-h timepoint suggests very low permeability or blood flow, but also could be related to previous preclinical findings suggestive of a blood–thymus barrier restricting access of high molecular weight particles from blood to T cells in thymus *(35,36)*. Additionally, while most CD4^+^ and CD8^+^ T cells in thymus are recently generated naïve T cells, preclinical studies in rodents, lamb, and pigs suggest that a small fraction of thymic mature T cells are immigrants from periphery *(37)* and therefore, migration of these mature CD8^+^ T cells, previously labeled with the radiotracer, back to thymus could further explain the thymus uptake only at the 48-h timepoint.

This pilot study had some limitations, particularly with a small number of participants. CD8 targeted imaging with ^89^Zr-Df-Crefmirlimab is in general limited to targeting all CD8^+^ cell types ‒ with no specificity to CD8^+^ T cells or their subsets ‒ and can include effects from changes both in CD8 expression and CD8^+^ cell population. Furthermore, it cannot distinguish between the relatively small percentage of antigen-specific CD8^+^ T cells, which are directly affected by the viral infection and its recovery process, and the vast majority of CD8^+^ T cells that do not recognize the antigen, but which may have been non-specifically activated due to the host inflammatory response through bystander activation. Future developments of kinetic models best describing the kinetics of ^89^Zr-Df-Crefmirlimab should include separate cell trafficking pathways in the model and should be accompanied by longitudinal blood sample collection. Separate studies would be required to quantify the probabilities of ^89^Zr dissociation from the minibody, observing ^89^Zr-labelled metabolites of the tracer, unbinding of the minibody from CD8 receptors, and exocytosis of an internalized minibody tracer in different tissues.

Lastly, dynamic immunoPET imaging is currently the only available non-invasive technology that can provide *in vivo* insight into whole-body CD8^+^ T cell distribution and trafficking in human subjects, and although based on a very small group of subjects, appears to offer higher sensitivity than the peripheral blood assays for studying CD8^+^ T cell physiology in individual subjects. The specificity of this and other immunoPET tracers under development, combined with the high detection sensitivity of total-body PET now provides a new platform for non-invasively and longitudinally studying the immune response and memory in all organs of the body in individual subjects in processes that challenge or stimulate the cell-mediated immune system. This includes, but is not limited to, cancer, infectious disease, autoimmune disease, and transplant patients and can be used for prognosis, as well as therapeutic and vaccine developments.

## MATERIALS AND METHODS

A pilot total-body PET imaging study with ^89^Zr-Df-Crefmirlimab was performed. The protocol was approved by the Institutional Review Board and all participants provided written informed consent.

### Study Design

The study consisted of two groups, including patients recovering from COVID-19 and healthy control subjects. The COVID-19 convalescent patients had a previous mild or moderate symptomatic infection and were not hospitalized. They all had positive identification of SARS-CoV-2 nucleic acids by a polymerase chain reaction (PCR) assay or SARS-CoV-2 nucleocapsid protein antigen identification at the time of diagnosis. The exclusion criteria for COVID-19 patients were subjects with serious comorbidities, history of splenic disorders or splenectomy, or use of medications that may impact T cells. Healthy controls gave no history of cancer or autoimmune disease within the last 5 years, no history of immune modulating therapy, no viral infection currently or within the 4 weeks prior to the study, and no history of COVID-19 infection, which was confirmed by negative detection of IgG antibodies against the nucleocapsid protein of SARS-CoV-2 in subjects vaccinated against COVID-19 and negative detection of IgG antibodies to the spike protein (S1/S2) of SARS-CoV-2 in unvaccinated subjects. A negative SARS-CoV-2 nucleic acids finding by a PCR assay was required prior to the first imaging visit for all subjects in both groups.

### Radiotracer Formulation and Administration

Crefmirlimab-berdoxam was radiolabeled with ^89^Zr at either Optimal Tracers, CA, USA or Memorial Sloan Kettering Cancer Center, NY, USA. Each patient dose contained a ∼1.5 mg mass dose of Crefmirlimab-berdoxam anti-CD8 minibody (ImaginAb, Inc., USA). The radiochemical purity determined by instant thin-layer chromatography was more than 95% in all cases. An ∼18.5 MBq (0.5 mCi) dose of ^89^Zr-Df-Crefmirlimab was infused intravenously over 5‒10 min using a syringe pump, followed by clearance with 30 cc of normal saline. Estimated effective radiation dose was 12 mSv. No pre-medications were administered, and vital signs were recorded pre-infusion, post-infusion, and at the end of imaging.

### PET/CT Imaging

Subjects had total-body PET/CT scans on the uEXPLORER scanner (United Imaging Healthcare, Shanghai, China) at three timepoints, including a 90-min PET dynamic scan starting immediately prior to the infusion, followed by two 60-min PET scans at 6 h and 48 h p.i. A low-dose CT scan (dose modulated, max 50 average mAs, 140 kVp, 10 mSv estimated effective radiation dose) was acquired prior to the first PET scan of each subject and ultra-low-dose CT scans (dose modulated, max 6 average mAs, 140 kVp, 1 mSv estimated effective radiation dose) were acquired prior to the later timepoint PET scans. The COVID-19 convalescent patients were first scanned within 8 weeks from onset of their symptoms and returned after 4±1 months for a second set of PET/CT scans.

PET images were reconstructed using the vendor’s image reconstruction software, which used an iterative time-of-flight ordered-subset expectation maximization (OS-EM) algorithm, with a reconstruction field-of-view of 60 cm and 4 iterations (20 subsets). A first set of images were reconstructed at a high spatial resolution, using a 512 × 512 matrix with 1.172 mm isotropic voxels and point spread function (PSF) modelling, using 60 min of the listmode data from each timepoint (30-90 min from the dynamic scans) and were used for visualization and localization of the volumes of interest (VOIs), particularly for smaller structures such as lymph nodes and vertebrae. A second set of images were reconstructed at a lower spatial resolution, using a 256 × 256 matrix with 2.344-mm isotropic voxels and no PSF modelling, which were used for data analysis in all organs. The dynamic datasets were reconstructed using the latter setting and 6 × 60 s, 16 × 30 s, 2 × 60 s, 12 × 120 s, and 10 × 300 s frames were generated. All corrections recommended by the manufacturer were applied and no post-reconstruction smoothing filters were used.

### Image Analysis

PET/CT images from 60-min reconstructions of all timepoints were first visualized in AMIDE medical image analysis software *(38)*. A qualitative assessment of the tracer distribution was performed at each time point and ∼100 spherical VOIs were drawn on each image over spleen, bone marrow (vertebrae, sacrum, and ilium), liver, lungs, thymus (when visible in PET), LV blood pool, right ventricle (RV) blood pool, head and neck lymph nodes, palatine tonsils, cerebrum, cerebellum, and nasal cavity mucosa. For large organs, several small VOIs were placed on the organ, excluding regions that may have been affected by motion and regions in proximity of blood vessels as much as possible. The coordinates and dimensions of the VOIs were transferred to MATLAB R2021b (The MathWorks, Inc., Natick, MA, USA) and image analysis was performed using an in-house developed code package. A segmented organ map was created for each dataset by first, calculating the mean and standard deviation of PET voxel values covered by the initial set of small VOIs for each organ; and subsequently, increasing the VOI diameters and assigning an organ index to the voxels with PET values within the mean ± standard deviation of the organ and CT values within a predefined Hounsfield unit range specific to the organ. The SUV ‒ defined as the ratio of image-derived activity concentration (decay corrected to the injection time) to the administered dose divided by the body weight ‒ was expressed as SUVmean for all organs, calculated from the mean of all voxels assigned to the organ; except for the lymph nodes, for which SUVpeak was calculated for each lymph node individually. SUVpeak was defined as the mean value the eight hottest voxels within the VOI, equivalent of ∼0.1 mL volume. Air fraction correction in the lungs was investigated, using the co-registered CT image values. Thymus fatty degeneration was evaluated in all subjects using the low-dose CT images, with four-point scores: Score 0 representing complete fatty replacement of the thymus and Score 3 representing a solid thymic gland with predominantly soft-tissue attenuation *(34)*. The TAC of image-derived LV blood pool activity concentration was plotted for each dataset and fitted with a tri-exponential function to derive the whole-blood clearance rate.

### Kinetic Modelling

TBR was calculated from the ratio of the tissue activity concentration in each organ (*C*_*T*_) to the whole-blood activity concentration (*C*_*p*_) at any given time. It should be noted that the subscript *p* in *C*_*p*_ conventionally represents the plasma activity concentration, whereas in this study image-derived whole-blood activity concentration from the LV blood pool was used for all organs, except for the lungs, for which the RV blood pool was used. Patlak graphs *(39)* were generated by plotting the TBR vs. normalized time defined as 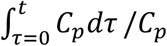, in which tri-exponential fitted whole-blood TACs were used for integration. Three conventional compartmental tracer kinetic models were fitted on each TAC to investigate feasibility of ^89^Zr-immuno PET kinetic modeling and to assess its reliability in representing the underlaying biology in different organs. This included the one-tissue compartmental model with three fitting microparameters (1T3P) of (*v*_*b*_, *K*_1_, *k*_2_), two-tissue compartmental model with four fitting microparameters (2T4P) of (*v*_*b*_, *K*_1_, *k*_2_, *k*_3_), and two-tissue compartmental model with five fitting microparameters (2T5P) of (*v*_*b*_, *K*_1_, *k*_2_, *k*_3_, *k*_4_) (Fig. S21). Model fitting was performed in all cases using all available datapoints up to 49-h p.i.. The Levenberg-Marquardt algorithm was used for nonlinear least squares fitting, using nonuniform weighting factors defined for each time frame based on the frame duration and decay factor. Due to higher uncertainties in the two late-timepoints at 6-h and 48-h p.i. ‒ particularly due to significantly higher statistical noise in the input function measurement as a result of blood clearance ‒ and availability of just a single data point at each late timepoint, the weighting factors were reduced by a factor of 10 for the two late timepoints. The AIC with a correction for small sample sizes was used as an estimate of the prediction error to choose the model best fitting the data. In case of the 2T model, *K*_*i*_ was calculated as a macroparameter representing the net influx rate of the tracer, defined as *K*_*i*_ = *K*_1_*k*_3_/(*k*_2_ + *k*_3_). To determine whether model parameters can be accurately estimated in presence of noise, practical identifiability analysis was performed in lungs, spleen, bone marrow (sacrum and ilium selected as regions less prone to motion), tonsils, and selected occipital lymph nodes, including calculation of normalized sensitivity curves, correlation matrix, bias, standard deviation, and root mean square error (RMSE) of the microparameters, as previously described *(40)*. The scaling factor used for modeling the TAC noise was calculated for each subject separately by comparing the measured TAC to the modeled TAC, and 100 TACs were simulated with the noise model in each case to calculate the bias and standard deviation of the microparameter estimates.

### Peripheral Blood Assays

Prior to the radiotracer infusion, a ∼20 mL whole-blood sample was drawn intravenously from each subject into vacutainer tubes containing ethylenediaminetetraacetic acid (EDTA). Peripheral blood mononuclear cells (PBMC) were isolated from whole blood by Ficoll-Hypaque (Cytiva, Marlborough, MA, USA) density gradient centrifugation and red blood cell lysis was performed using ACK lysis buffer (Gibco/ThermoFisher Scientific, Waltham, MA, USA). PBMC were viably cryopreserved in fetal calf serum (FCS) with 10% dimethyl sulfoxide (DMSO), stored at - 140° C in liquid nitrogen (LN2).

At the conclusion of the study, PBMC were thawed and rested overnight in R15 (RPMI-1640 supplemented with 15% FCS, 100 U/mL penicillin, 100 mg/mL streptomycin, and 2 mM L-glutamine) at 37°C, 5% CO2. Immunophenotyping was performed by first staining the PBMC for dead cells using a fixable amine-reactive viability dye, followed by an extracellular stain using the fluorescently conjugated monoclonal antibodies listed in Table S5 in the presence of brilliant stain buffer (BD Bioscience) to reduce aggregate formation. Cells were then fixed in 1% formaldehyde and flow cytometry was performed within 24 h. CD8^+^ and CD4^+^ T cells identified from viable CD3^+^ cells were further delineated based on CD45RA and CCR7 expression into memory subsets: central memory (TCM), effector memory (TEM), terminally differentiated effector memory (TEMRA), or naïve T cells (Fig. S22). T cell activation was investigated based on CD56 expression and co-expression of HLA-DR and CD38, and T cell exhaustion was assessed by PD-1 expression (Fig. S23). Treg cells (CD4^+^, CD25^+^, CD127^-^) and MAIT cells (CD161^+^, Vα7.2^+^) were gated, and lastly, NK cells were gated based on CD56 and CD16 expression and B cells were gated based on CD19 (Fig. S24).

PBMC were also assessed for responsiveness to SARS-CoV-2 peptides by intracellular cytokine staining, as previously described *(41,42)*, using the flow cytometry panel described in Table S6. After overnight resting, PBMC were incubated in R15 in the presence of stimulation cocktail, including CD107a-PE-Cy5 (to measure the degranulation response), unlabeled CD28 and CD49d costimulatory antibodies, and the protein transport inhibitors, brefeldin A (MilliporeSigma, St. Louis, MO, USA) and Golgi Stop (monensin, BD Biosciences, Franklin Lakes, NJ, USA) Cells were stimulated with SARS-CoV2 peptide pools (15mers overlapping by 11) spanning the spike and nucleocapsid proteins (JPT Peptides, Berlin, DE) at a concentration of 3.5 μg/mL for 5 hours at 37°C, 5% CO2. DMSO (peptide carrier) served as a negative control and staphylococcal enterotoxin B (SEB; 5 μg/mL) served as a positive control. Following stimulation, cells were stained for viability and extracellular markers. They were then fixed in 4% formaldehyde, permeabilized using BD FACS Perm 2 (BD Biosciences), and intracellular staining was performed. Brilliant stain buffer was used during the extracellular and intracellular staining steps to minimize aggregate formation. Samples were then fixed in 1% formaldehyde and flow cytometry was performed on the next day. Detection of intracellular effector molecules (IFNγ, IL-2, TNF-α, MIP-1β, and granzyme B) as well as the degranulation (CD107a) response were measured in memory CD8^+^ and CD4^+^ T cells (Fig. S25). The data were analyzed to look at the total response, each of the individual responses, and the polyfunctional response (i.e., production of combinations of the analytes above). A previously described statistical algorithm *(43)*, based on the total number of collected events (memory CD8^+^ or CD4^+^) utilizing a Poisson distribution was used to determine if stimulated responses differed significantly from unstimulated samples to perform background subtraction.

In all cases, flow cytometry was performed with a 5-laser, 40-color Aurora spectral cytometer (Cytek, Fremont, CA, USA) and the data were analyzed using FlowJo software version 10.8.1 (BD Biosciences). Polyfunctionality was mapped in SPICE software version 6.1 *(44)*.

### Statistical Analysis

Hypothesis testing comparing the two sets of scans from the COVID-19 group to the control group were performed on all datasets in GraphPad Prism version 9.5, using a two-tailed unpaired Mann-Whitney U test. P values <0.05 were set to determine statistical significance.

## Data Availability

All data associated with this study are present in the paper or the Supplementary Materials. The analysis codes, raw analysis data, and anonymized image data used in this article will be made available by the authors upon request, to any researcher for purposes of reproducing or extending the analysis.

## LIST OF SUPPLEMENTARY MATERIALS

Fig. S1 to S25

Tables S1 to S6

## ACKNOWLEDGMENTS

The authors would like to thank Dr. Elizabeth Li, Yiran Wang, and Dr. Benjamin Spencer for their technical and scientific support during this project, and Dr. Guobao Wang, Dr. Adriaan Lammertsma, Dr. Roger Gunn, and Dr. Maani Archang for the fruitful discussions and their valuable feedback on this work. Furthermore, the authors thank Lynda Painting (research coordinator) and the team of technologists at EXPLORER Molecular Imaging Center for their assistance in the imaging study. Furthermore, the authors thank the UC Davis Flow Cytometry Shared Resource Laboratory staff members, Ms. Bridget McLaughlin and Mr. Jonathan van Dyke, for their assistance with the flow cytometry.

## FUNDING

National Institutes of Health grant R01CA206187 (RDB, SRC)

National Institutes of Health grant R35CA197608 (SRC)

ImaginAb, Inc., Inglewood, CA, USA

National Institutes of Health NCI grant P30 CA0933730 (PL)

National Institutes of Health NCRR grant S10 RR12964 (PL)

National Institutes of Health NCRR grant S10 RR026825 (JN)

National Institutes of Health NCRR grant S10 OD018223-01A1 (BLS)

James B. Pendleton Charitable Trust (awarded to a committee led by BLS)

## AUTHOR CONTRIBUTIONS

Conceptualization: NO, TJ, PMP, IW, SRC

Methodology: NO, TJ, PMP, ALF, JL, YGA, FS, SHC, RDB, BLS, IW, SRC

Investigation: NO, TJ, PMP, ALF, JL, YGA, BLS, SRC

Visualization: NO, ALF

Funding acquisition: RDB, IW, SRC

Project administration: FS, SHC, RDB, BLS, IW, SRC

Supervision: TJ, PMP, BLS, IW, SRC

Writing – original draft: NO, ALF

Writing – review & editing: NO, TJ, PMP, ALF, JL, YGA, FS, SHC, KS, RDB, BLS, IW, SRC

## COMPETING INTERESTS

RDB and SRC are principal investigators on a grant funded by United Imaging Healthcare. UC Davis has a research agreement and a sales-based revenue sharing agreement with United Imaging Healthcare. This research was supported by ImaginAb. IW serves as CEO of ImaginAb. KS serves as VP of Medical Affairs at ImaginAb.

## Notes

### Competing Interest Statement

RD Badawi and SR Cherry are principal investigators on a grant funded by United Imaging Healthcare. UC Davis has a research agreement and a sales-based revenue sharing agreement with United Imaging Healthcare. This research was supported by ImaginAb. I Wilson serves as CEO of ImaginAb. K Schmiedehausen serves as VP of Medical Affairs at ImaginAb.

### Clinical Trial

IND 153492

### Funding Statement

This study was funded by National Institutes of Health grants R01CA206187, R35CA197608, P30 CA0933730, S10 RR12964, S10 RR026825, S10 OD018223-01A1, as well as James B. Pendleton Charitable Trust and ImaginAb, Inc., Inglewood, CA, USA.

### Author Declarations

Ethics committee/IRB of University of California Davis gave ethical approval for this work.

