## Supplementary_Materials for "First-in-human immunoPET imaging of COVID-19 convalescent patients using dynamic total-body PET and a CD8-targeted minibody"

**Table S1. Complete demographics of the participants.** Vaccination and diagnosis timelines are included. Note: No patient recorded central nervous system or cardiac symptoms of COVID-19.

| Subject number | Study group | Sex | Age Range | BMI | Dynamic scan | Vaccination and diagnosis Timeline |
|---|---|---|---|---|---|---|
| 1 | COVID-19 | F | 51-55 | 38 | - | Infected with COVID-19 in March 2021 before vaccination and was sick for 3-4 weeks.<br>Symptoms: shortness of breath, abdominal pain, nausea, mild headache<br>First PET scan was 11 days after the first vaccination dose and second PET scan was 3 months after the second vaccination dose.<br>Past medical history: Cushing disease (pituitary removed in 2012), diabetes, hyperlipidemia, and hypothyroid |
| 2 | COVID-19 | F | 26-30 | 29 | 90 min | Infected with COVID-19 in January 2021 (asymptomatic), 13 days after the first vaccination dose. Infected with COVID-19 again in April 2021, 3 months after the second dose of vaccination and was sick for 2 weeks.<br>Symptoms: loss of taste and smell, fatigue, difficulty breathing, headaches, nausea, congestion, sore throat, runny nose, extreme stomach pain after eating, lightheadedness, extreme exhaustion, shortness of breath, extreme body aches<br>First PET scan was 4 months after the second vaccination dose and second PET scan was 8.5 months after the second vaccination dose.<br>Past medical history: gastric bypass, iron deficiency anemia, back pain with radiculopathy, laminectomy |
| 3 | COVID-19 | F | 46-50 | 43 | - | Infected with COVID-19 in May 2021, 3.5 months after the second vaccination dose and was sick for 1 week.<br>Symptoms: chronic cough, headache, sore throat, fever, chills, abdominal pain, congestion, body aches, dysuria, muscle pain<br>First PET scan was 4.5 months after the second vaccination dose and second PET scan was 8.5 months after the second vaccination dose.<br>Past medical history: cholecystectomy, sleeve gastrectomy, tubal ligation |
| 4 | COVID-19 | F | 26-30 | 35 | 90 min | Infected with COVID-19 in February 2022, 3 months after the third vaccination dose and was sick mildly for 5-6 days.<br>Symptoms: cough, diarrhea, fatigue, fever, slight headache, sore throat<br>First PET scan was 4.5 months after the second vaccination dose and second PET scan was 7 months after the third vaccination dose.<br>Past medical history: none |
| 5 | COVID-19 | F | 31-35 | 20 | 90 min | Infected with COVID-19 in March 2022, 8 months after the second vaccination dose and was mildly sick for 3 days.<br>Symptoms: cough, fatigue, congestion, runny nose<br>First PET scan was 9.5 months after the second vaccination dose and second PET scan was 4 months after the third vaccination dose.<br>Past medical history: cervical polypectomy. |
| 6 | Control | M | 21-25 | 21 | 90 min | First PET scan was 6 months after the second vaccination dose.<br>Past medical history: not available. |
| 7 | Control | M | 46-50 | 25 | 65 min | First PET scan was 6 months after the second vaccination dose.<br>Past medical history: femoral bone fracture, hypertension. |
| 8 | Control | F | 56-60 | 31 | 90 min | Not vaccinated.<br>Past medical history: not available. |

**Table S2. Microparameters of model fits in all organs-of-interest of individual subjects.** Microparameters of AIC-preferred model fit results for lungs, spleen, sacrum, ilium, tonsils, and occipital lymph nodes shown for all subjects with dynamic scans. $K_1$ is shown in $ml_{plasma}/min/ml_{tissue}$, and $k_2$, $k_3$, and $k_4$ are shown in 1/min. $Ki$ is calculated by $\frac{K_1 k_3}{k_2+k_3}$ and $V_T$ is calculated by $\frac{K_1}{k_2}(1+\frac{k_3}{k_4})$.

**Lungs - 2T5P**

| | $v_b$ | $K_1$ | $k_2$ | $k_3$ | $k_4$ | $K_i$ | $V_T$ |
|---|---|---|---|---|---|---|---|
| Sub02 - Baseline | 0.22 | 0.02 | 0.16 | 0.004 | 0.0013 | 0.00054 | 0.56 |
| Sub02 – Follow-up | 0.24 | 0.03 | 0.18 | 0.017 | 0.0074 | 0.00239 | 0.50 |
| Sub04 - Baseline | 0.17 | 0.00 | 1.00 | 0.050 | 0.0006 | 0.00018 | 0.31 |
| Sub04 - Follow-up | 0.14 | 0.05 | 0.44 | 0.001 | 0.0003 | 0.00014 | 0.56 |
| Sub05 - Baseline | 0.17 | 0.01 | 0.58 | 0.003 | 0.0000 | 0.00005 | - |
| Sub05 - Follow-up | 0.16 | 0.00 | 0.04 | 0.009 | 0.0005 | 0.00012 | 0.30 |
| Sub06 | 0.13 | 0.02 | 0.50 | 0.001 | 0.0000 | 0.00004 | - |
| Sub07 | 0.23 | 0.01 | 0.52 | 0.000 | 0.0029 | 0.00000 | 0.01 |
| Sub08 | 0.18 | 0.02 | 0.32 | 0.002 | 0.0001 | 0.00012 | 1.28 |

**Spleen - 2T5P**

| | $v_b$ | $K_1$ | $k_2$ | $k_3$ | $k_4$ | $K_i$ | $V_T$ |
|---|---|---|---|---|---|---|---|
| Sub02 - Baseline | 0.40 | 1.48 | 0.04 | 0.0040 | 0.0015 | 0.12 | 124.4 |
| Sub02 – Follow-up | 0.40 | 1.43 | 0.04 | 0.0049 | 0.0021 | 0.17 | 128.5 |
| Sub04 - Baseline | 0.40 | 0.88 | 0.06 | 0.0037 | 0.0007 | 0.05 | 86.8 |
| Sub04 - Follow-up | 0.40 | 1.05 | 0.06 | 0.0047 | 0.0009 | 0.07 | 101.8 |
| Sub05 - Baseline | 0.40 | 1.08 | 0.08 | 0.0028 | 0.0005 | 0.04 | 87.0 |
| Sub05 - Follow-up | 0.40 | 0.88 | 0.07 | 0.0030 | 0.0005 | 0.03 | 88.2 |
| Sub06 | 0.40 | 0.84 | 0.07 | 0.0026 | 0.0004 | 0.03 | 87.0 |
| Sub07 | 0.40 | 0.85 | 0.07 | 0.0018 | 0.0003 | 0.02 | 99.0 |
| Sub08 | 0.40 | 1.09 | 0.06 | 0.0033 | 0.0007 | 0.05 | 97.6 |

**Sacrum - 2T5P**

| | $v_b$ | $K_1$ | $k_2$ | $k_3$ | $k_4$ | $K_i$ | $V_T$ |
|---|---|---|---|---|---|---|---|
| Sub02 - Baseline | 0.06 | 0.08 | 0.02 | 0.0041 | 0.0026 | 0.017 | 13.1 |
| Sub02 – Follow-up | 0.08 | 0.09 | 0.01 | 0.0033 | 0.0036 | 0.022 | 16.9 |
| Sub04 - Baseline | 0.06 | 0.04 | 0.02 | 0.0027 | 0.0008 | 0.004 | 8.3 |
| Sub04 - Follow-up | 0.02 | 0.04 | 0.02 | 0.0046 | 0.0012 | 0.007 | 8.9 |
| Sub05 - Baseline | 0.02 | 0.09 | 0.04 | 0.0015 | 0.0005 | 0.003 | 8.9 |
| Sub05 - Follow-up | 0.05 | 0.08 | 0.04 | 0.0015 | 0.0004 | 0.003 | 10.8 |
| Sub06 | 0.04 | 0.05 | 0.04 | 0.0019 | 0.0005 | 0.002 | 5.5 |
| Sub07 | 0.03 | 0.05 | 0.03 | 0.0019 | 0.0003 | 0.003 | 11.8 |
| Sub08 | 0.03 | 0.03 | 0.02 | 0.0008 | 0.0001 | 0.001 | 9.9 |

**Ilium - 2T5P**

| | $v_b$ | $K_1$ | $k_2$ | $k_3$ | $k_4$ | $K_i$ | $V_T$ |
|---|---|---|---|---|---|---|---|
| Sub02 - Baseline | 0.05 | 0.08 | 0.02 | 0.0039 | 0.0026 | 0.015 | 12.0 |
| Sub02 – Follow-up | 0.08 | 0.08 | 0.01 | 0.0032 | 0.0038 | 0.020 | 16.0 |
| Sub04 - Baseline | 0.00 | 0.05 | 0.02 | 0.0025 | 0.0008 | 0.005 | 8.3 |
| Sub04 - Follow-up | 0.03 | 0.04 | 0.01 | 0.0017 | 0.0007 | 0.005 | 10.5 |
| Sub05 - Baseline | 0.02 | 0.08 | 0.04 | 0.0014 | 0.0004 | 0.003 | 8.9 |
| Sub05 - Follow-up | 0.03 | 0.07 | 0.04 | 0.0018 | 0.0005 | 0.003 | 9.2 |
| Sub06 | 0.02 | 0.04 | 0.04 | 0.0015 | 0.0005 | 0.002 | 5.1 |
| Sub07 | 0.04 | 0.04 | 0.02 | 0.0015 | 0.0002 | 0.002 | 10.8 |
| Sub08 | 0.00 | 0.03 | 0.02 | 0.0013 | 0.0005 | 0.002 | 5.1 |

**Tonsils - 1T3P**

| | $v_b$ | $K_1$ | $k_2$ | $K_1/k_2$ |
|---|---|---|---|---|
| Sub02 - Baseline | 0.00 | 0.014 | 0.00053 | 25.9 |
| Sub02 – Follow-up | 0.00 | 0.012 | 0.00036 | 32.6 |
| Sub04 - Baseline | 0.00 | 0.004 | 0.00018 | 22.3 |
| Sub04 - Follow-up | 0.06 | 0.005 | 0.00012 | 42.0 |
| Sub05 - Baseline | 0.04 | 0.005 | 0.00023 | 22.7 |
| Sub05 - Follow-up | 0.03 | 0.004 | 0.00018 | 22.5 |
| Sub06 | 0.05 | 0.004 | 0.00014 | 30.2 |
| Sub07 | 0.05 | 0.001 | 0.00000 | - |
| Sub08 | 0.05 | 0.002 | 0.00000 | - |

**Occipital Lymph Nodes - 1T3P**

| | $v_b$ | $K_1$ | $k_2$ | $K_1/k_2$ |
|---|---|---|---|---|
| Sub02 - Baseline | 0.02 | 0.0009 | 0.00040 | 2.2 |
| Sub02 – Follow-up | 0.02 | 0.0010 | 0.00092 | 1.0 |
| Sub04 - Baseline | 0.05 | 0.0012 | 0.00000 | - |
| Sub04 - Follow-up | 0.05 | 0.0017 | 0.00000 | - |
| Sub05 - Baseline | 0.05 | 0.0020 | 0.00000 | - |
| Sub05 - Follow-up | 0.05 | 0.0020 | 0.00000 | - |
| Sub06 | 0.06 | 0.0002 | 0.00019 | 1.2 |
| Sub07 | 0.14 | 0.0005 | 0.00000 | - |
| Sub08 | 0.00 | 0.0021 | 0.00024 | 8.9 |

**Table S3. Correlation matrix of model microparameters.** Correlation matrix of microparameters of the AIC-preferred model is presented for lungs, spleen, sacrum, ilium, tonsils, and occipital lymph nodes, showing mean and standard deviations calculated over all subjects with 90-min dynamic scans.

**Lungs**

| | $v_b$ | $K_1$ | $k_2$ | $k_3$ | $k_4$ |
|---|---|---|---|---|---|
| $v_b$ | 1.00 | -0.13 ± 0.64 | 0.30 ± 0.53 | -0.81 ± 0.20 | 0.71 ± 0.10 |
| $K_1$ | | 1.00 | -0.95 ± 0.05 | 0.51 ± 0.51 | -0.31 ± 0.43 |
| $k_2$ | | | 1.00 | -0.61 ± 0.46 | 0.39 ± 0.39 |
| $k_3$ | | | | 1.00 | -0.80 ± 0.08 |
| $k_4$ | | | | | 1.00 |

**Spleen**

| | $v_b$ | $K_1$ | $k_2$ | $k_3$ | $k_4$ |
|---|---|---|---|---|---|
| $v_b$ | 1.00 | -1.00 ± 0.00 | 0.96 ± 0.04 | -0.79 ± 0.06 | 0.33± 0.07 |
| $K_1$ | | 1.00 | -0.95 ± 0.04 | 0.76 ± 0.06 | -0.28 ± 0.07 |
| $k_2$ | | | 1.00 | -0.86 ± 0.02 | 0.43 ± 0.09 |
| $k_3$ | | | | 1.00 | 0.75 ± 0.05 |
| $k_4$ | | | | | 1.00 |

**Sacrum**

| | $v_b$ | $K_1$ | $k_2$ | $k_3$ | $k_4$ |
|---|---|---|---|---|---|
| $v_b$ | 1.00 | -0.86 ± 0.09 | 0.92 ± 0.05 | -0.81 ± 0.08 | 0.48 ± 0.06 |
| $K_1$ | | 1.00 | -0.91 ± 0.05 | 0.55 ± 0.16 | -0.12 ± 0.14 |
| $k_2$ | | | 1.00 | -0.75 ± 0.18 | 0.37 ± 0.25 |
| $k_3$ | | | | 1.00 | -0.81 ± 0.07 |
| $k_4$ | | | | | 1.00 |

**Ilium**

| | $v_b$ | $K_1$ | $k_2$ | $k_3$ | $k_4$ |
|---|---|---|---|---|---|
| $v_b$ | 1.00 | -0.86 ± 0.10 | 0.91 ± 0.05 | -0.79 ± 0.07 | 0.46 ± 0.07 |
| $K_1$ | | 1.00 | -0.91 ± 0.05 | 0.52 ± 0.09 | -0.08 ± 0.15 |
| $k_2$ | | | 1.00 | -0.74 ± 0.15 | 0.34 ± 0.25 |
| $k_3$ | | | | 1.00 | -0.81 ± 0.06 |
| $k_4$ | | | | | 1.00 |

**Tonsils**

| | $v_b$ | $K_1$ | $k_2$ |
|---|---|---|---|
| $v_b$ | 1.00 | -0.99 ± 0.00 | 0.85 ± 0.02 |
| $K_1$ | | 1.00 | -0.87 ± 0.03 |
| $k_2$ | | | 1.00 |

**Occipital Lymph Nodes**

| | $v_b$ | $K_1$ | $k_2$ |
|---|---|---|---|
| $v_b$ | 1.00 | -0.93 ± 0.07 | 0.79 ± 0.09 |
| $K_1$ | | 1.00 | -0.88± 0.03 |
| $k_2$ | | | 1.00 |

**Table S4. Estimated errors of model microparameters.** Bias (%), standard deviation (%), and RMSE (%) of the AIC-preferred model microparameters in lungs, spleen, sacrum, ilium, tonsils, and cervical lymph nodes, calculated from model fitting on 100 simulated TACs generated for each subject from the calculated noise model of the measured TAC. The results show the averaged values among all subjects with 90-min dynamic scans.

**Bias (%)**

| | Lungs | Spleen | Sacrum | Ilium | Tonsils | Occipital lymph nodes |
|---|---|---|---|---|---|---|
| $v_b$ | 2.9 | 0.7 | 1.0 | -0.4 | 1.6 | 7.9 |
| $K_1$ | -9.8 | 5.6 | -0.1 | -0.2 | 0.2 | 0.8 |
| $k_2$ | -9.0 | -0.1 | -0.2 | -0.5 | -0.7 | -1.9 |
| $k_3$ | 59.8 | 0.1 | -0.3 | -1.0 | | |
| $k_4$ | 26.2 | 0.1 | -0.6 | -1.2 | | |

**Standard deviation (%)**

| | Lungs | Spleen | Sacrum | Ilium | Tonsils | Occipital lymph nodes |
|---|---|---|---|---|---|---|
| $v_b$ | 6.7 | 34.3 | 29.0 | 40.5 | 17.2 | 18.7 |
| $K_1$ | 77.4 | 22.3 | 1.8 | 2.3 | 2.3 | 7.2 |
| $k_2$ | 63.9 | 4.4 | 4.7 | 6.2 | 7.7 | 22.4 |
| $k_3$ | 113.8 | 3.1 | 6.4 | 10.5 | | |
| $k_4$ | 197.3 | 2.7 | 7.0 | 7.0 | | |

**RMSE (%)**

| | Lungs | Spleen | Sacrum | Ilium | Tonsils | Occipital lymph nodes |
|---|---|---|---|---|---|---|
| $v_b$ | 7.5 | 34.3 | 29.2 | 40.4 | 17.3 | 20.8 |
| $K_1$ | 82.8 | 23.1 | 1.8 | 2.3 | 2.3 | 7.4 |
| $k_2$ | 67.7 | 4.4 | 4.7 | 6.2 | 8.9 | 22.4 |
| $k_3$ | 132.5 | 3.1 | 6.4 | 10.5 | | |
| $k_4$ | 200.4 | 2.7 | 7.0 | 7.1 | | |

**Table S5. Flow cytometry panel design used for cell immunophenotyping.** All antibodies and viability dyes were titrated to determine the optimal concentration for each assay.

| Antibody/Reagent | Clone | Fluorochrome | Producer | Significance |
|---|---|---|---|---|
| Viability | -- | Live/Dead Blue | ThermoFisher Scientific | Live/Dead |
| CD3 | UCHT1 | Alexa Fluor 532 | ThermoFisher Scientific | T cell marker |
| CD4 | RPA-T4 | APC-Fire750 | Biolegend | “Helper” T cells |
| CD8 | SK1 | BUV805 | BD Biosciences | “Cytotoxic” T cells |
| CD16 | 3G8 | BV480 | BD Biosciences | FcγRIII, NK |
| CD56 | HCD56 | BV650 | Biolegend | NK, activated CD8 |
| Vα7.2 | 3C10 | PE-Cy7 | Biolegend | MAIT |
| CD161 | DX12 | BV605 | BD Biosciences | NK, MAIT |
| CD25 | M-A251 | PE | Biolegend | IL-2R, Treg |
| CD127 | A019D5 | BV750 | Biolegend | IL-7Rα |
| CD19 | HIB19 | FITC | BD Biosciences | B-cells |
| CD45RA | HI100 | BUV395 | BD Biosciences | Naïve/TEMRA T cells |
| CCR7 | 3D12 | BV421 | BD Biosciences | Naïve/Central Memory T cells |
| CD38 | HIT2 | PE-CF594 | BD Biosciences | Activation marker (with HLA-DR) |
| HLA-DR | TU36 | PerCP-Cy5.5 | Biolegend | MHC-II; Activation marker (with CD38) |
| PD-1 | J105 | APC | ThermoFisher Scientific | T-cell exhaustion |

**Table S6. Flow cytometry panel design used for intracellular cytokine staining assays.** All antibodies and viability dyes were titrated to determine the optimal concentration for each assay.

| Antigen/Marker | Clone | Fluorochrome | | Significance |
|---|---|---|---|---|
| Viability | -- | Live/Dead Aqua | ThermoFisher Scientific | |
| CD3 | UCHT1 | Alexa Fluor 532 | ThermoFisher Scientific | T cell marker |
| CD4 | SFCI12T4D11 | PE-Cy7 | Beckman Coulter | “Helper” T-cells |
| CD8 | SK1 | BUV805 | BD Biosciences | “Cytotoxic” T-cells |
| CD45RA | HI100 | BUV395 | BD Biosciences | Naïve/TEMRA |
| CCR7 | 3D12 | BV421 | BD Biosciences | Naïve/Central Memory |
| CD107a | H4A3 | PE-Cy5 | BD Biosciences | Cytotoxic granules |
| CD28 | L293 | -- | BD Biosciences | Costimulatory |
| CD49d | L25 | -- | BD Biosciences | Costimulatory |
| IFNγ | 25723.11 | APC | BD Biosciences | Proinflammatory |
| IL2 | MQ1-17H12 | BV605 | BD Biosciences | T-cell growth |
| MIP-1β | D21-1351 | PE | BD Biosciences | Chemotactic |
| TNFα | MAb11 | BV711 | Biolegend | Proinflammatory |
| Granzyme B | GB11 | Alexa Fluor700 | BD Biosciences | Cytotoxic granules |

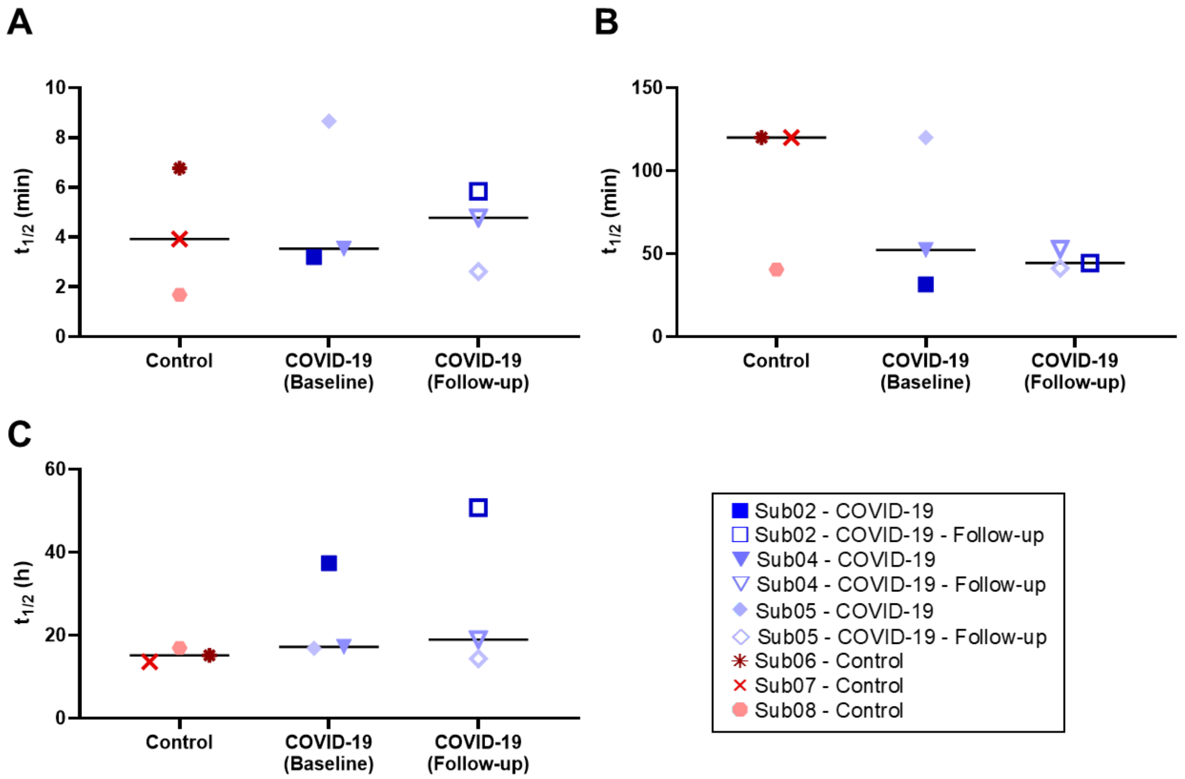


**Fig. S1. Whole-blood clearance biological half-lives.** Biological half-lives derived from triexponential fitting on the whole-blood TACs of 6 subjects (9 scans), including (**A**) the initial, (**B**) the intermediate, and (**C**) the terminal elimination phases. Image-derived LV blood pool was used for obtaining the whole-blood TACs. With a very low uptake in the myocardium, negligible effects from myocardium spill-over on LV blood pool were observed.

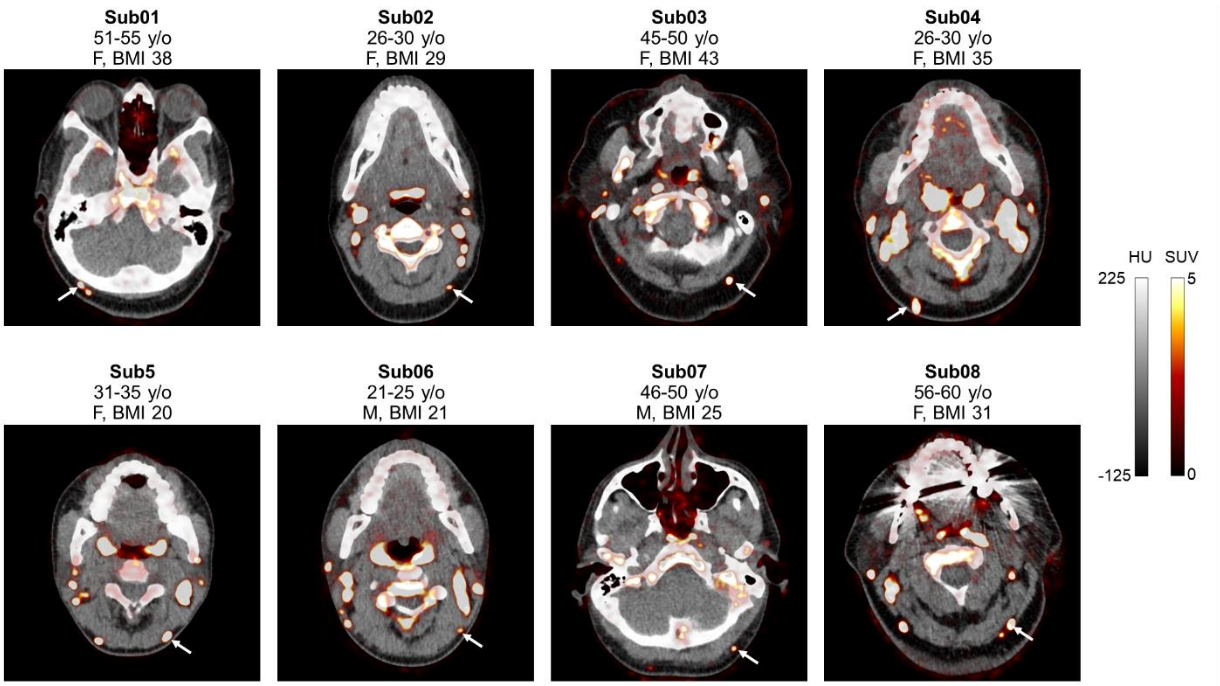


**Fig. S2. PET/CT images of selected occipital lymph nodes.** Transverse PET/CT slices through representative occipital lymph nodes selected in each subject (marked with arrows), which have been used for kinetic modeling. For COVID-19 patients (Sub01 to Sub05), only the baseline scans are shown.

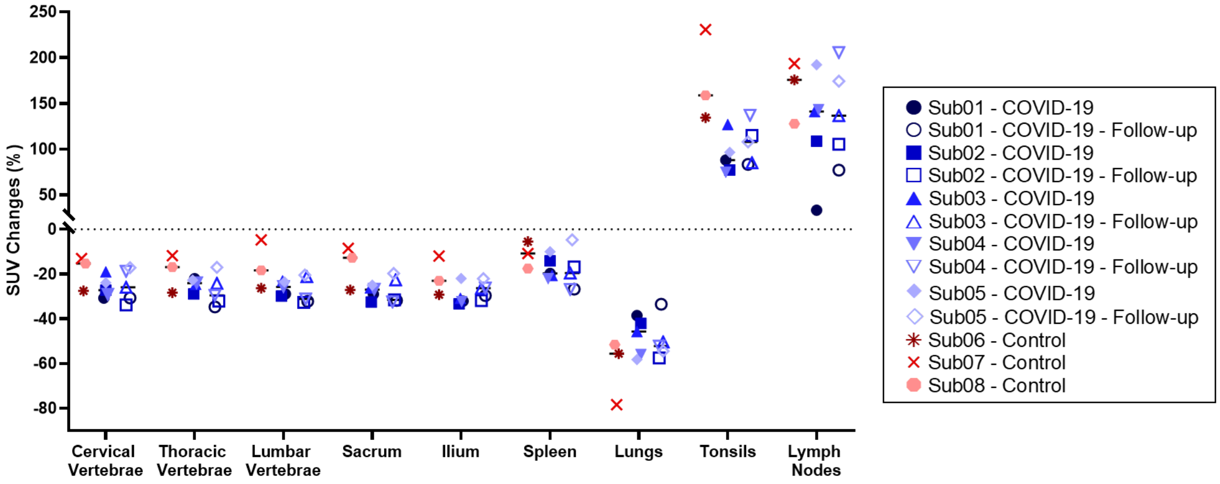


**Fig. S3. Percentage changes of SUV at the 48-h relative to the 6-h timepoint.** Percentage changes of SUV at the 48-h relative to the 6-h timepoint show similar trends in all subjects, in different regions of bone marrow (vertebrae, sacrum, and ilium), spleen, tonsils, and lymph nodes. The percentage changes of SUV in the lymph nodes represent the average SUV changes in 11–22 head and neck lymph nodes for each subject.

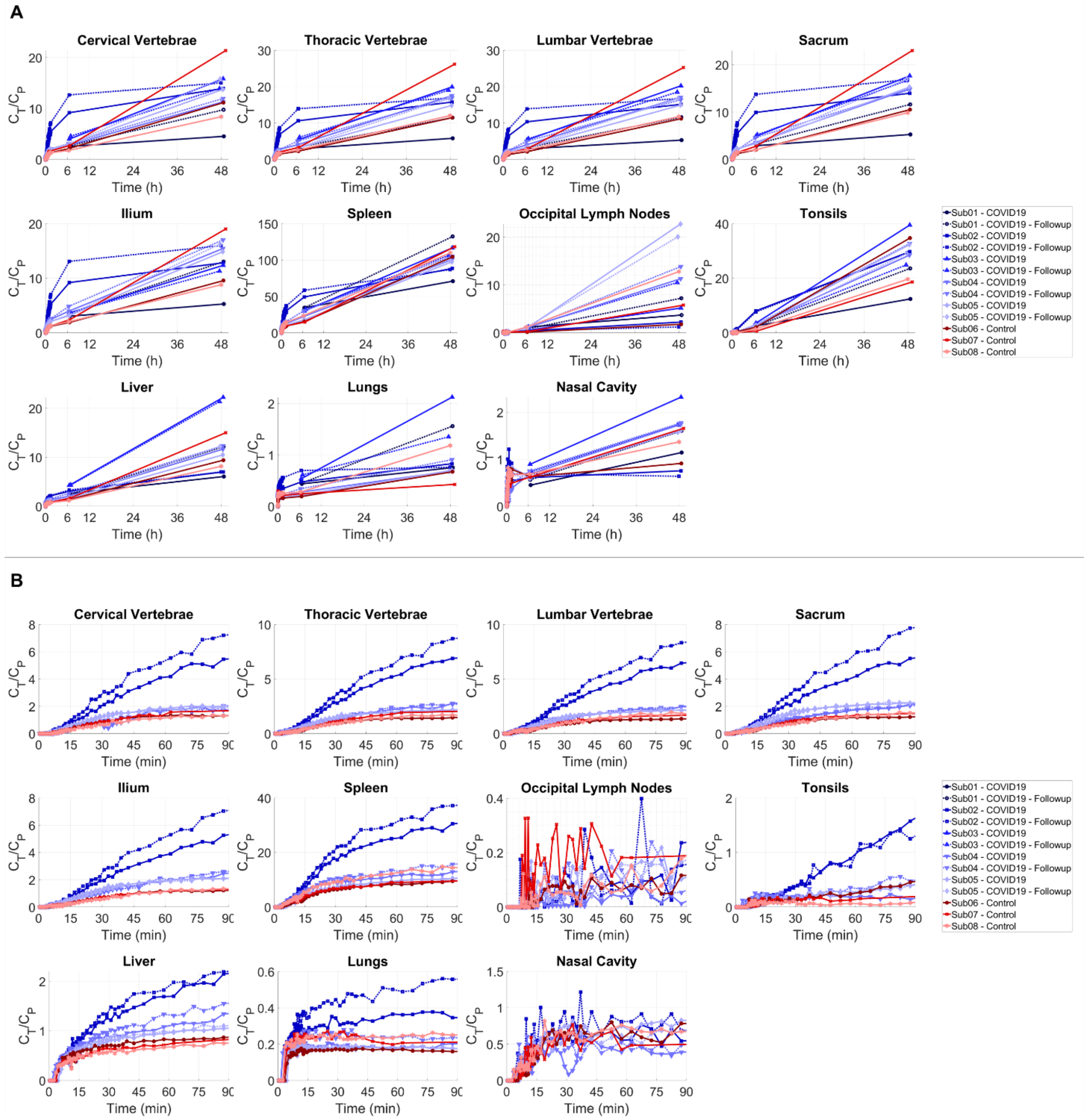


**Fig. S4. Decay-corrected TBR curves in different organs-of-interest.** TBRs are shown as a function of time for bone marrow, spleen, occipital lymph nodes, tonsils, liver, lungs, and nasal cavity (**A**) during the 48-h of the imaging study and (**B**) during the first 90-min after tracer administration for all subjects.

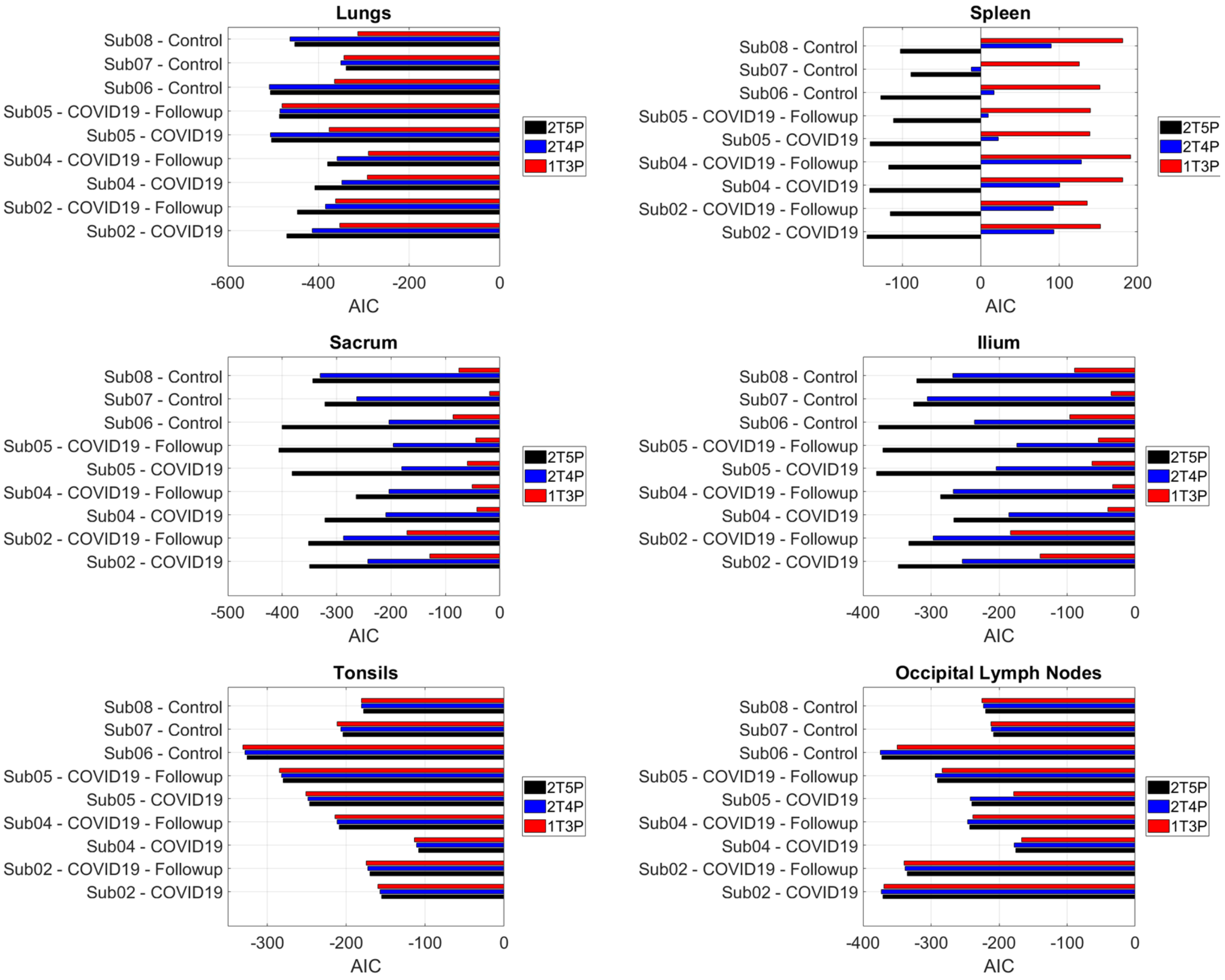


**Fig. S5. AIC model selection results in all organs-of-interest.** AIC values of 1T3P, 2T4P, and 2T5P model fittings performed on the lungs, spleen, bone marrow (sacrum and ilium), tonsils, and selected occipital lymph nodes of all subjects.

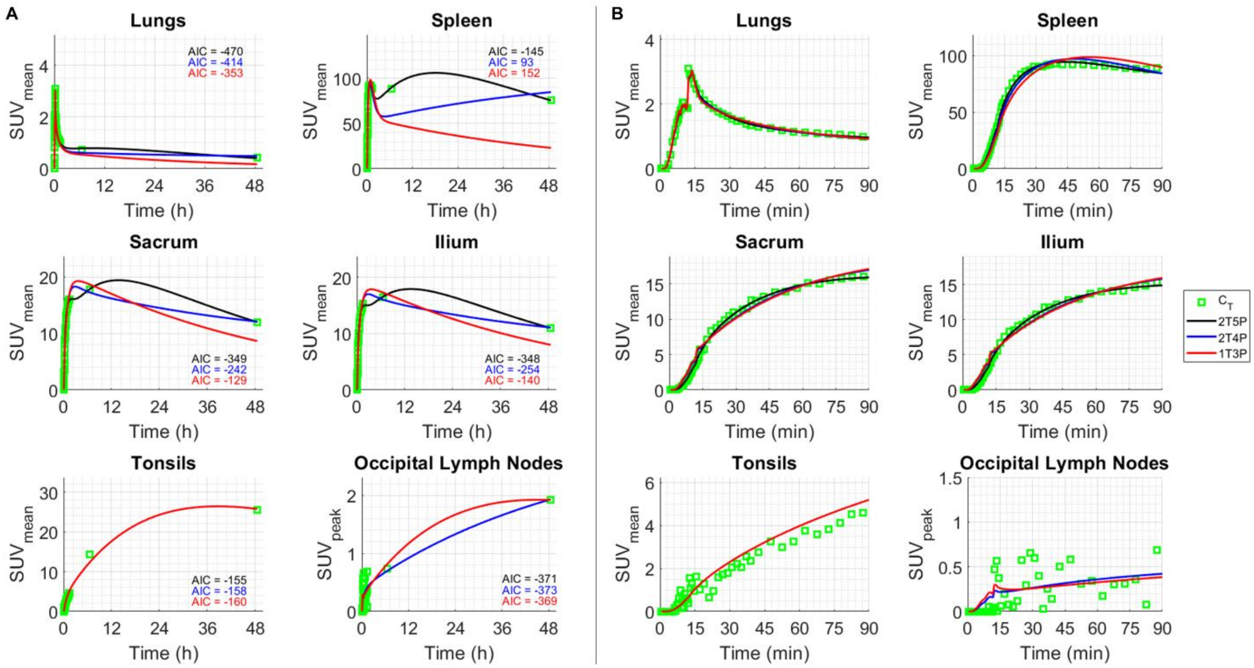


**Fig. S6. Examples of model fits in a COVID-19 convalescent subjects.** Results of 1T3P, 2T4P, and 2T5P model fittings performed on the lungs, spleen, bone marrow (sacrum and ilium), tonsils, and selected occipital lymph nodes of the baseline scans of an example COVID-19 patient (Sub02) shown for (**A**) the 0–48 h and (**B**) zoomed on the 0–90 min.

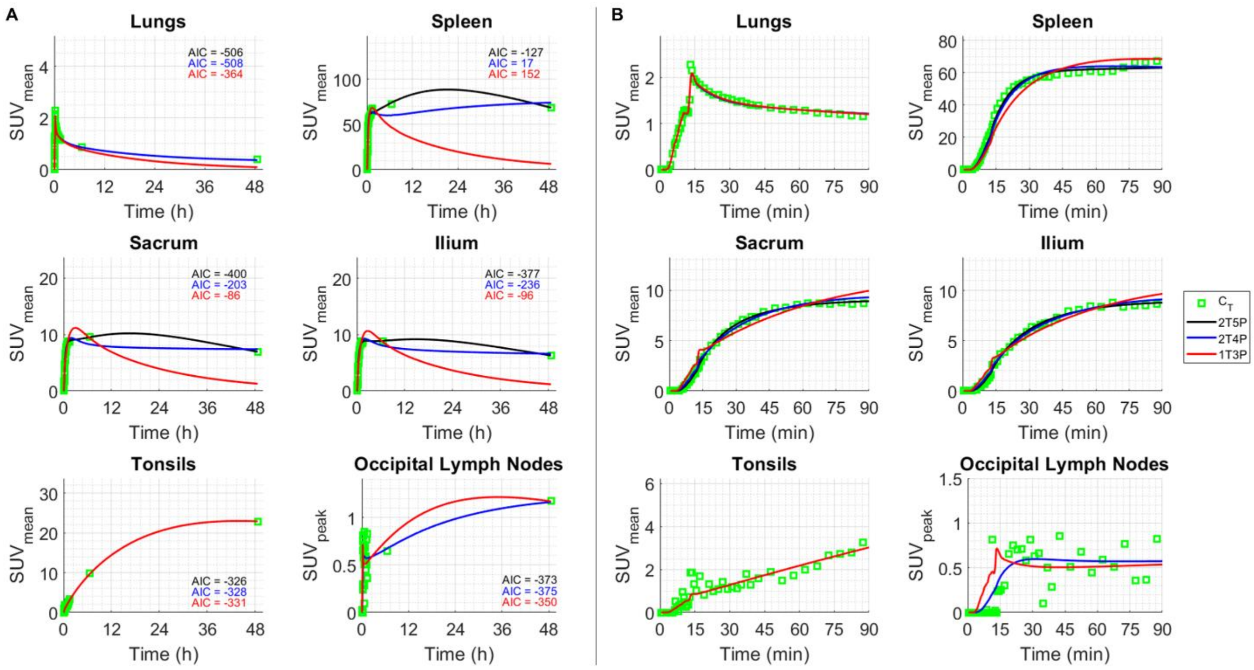


**Fig. S7. Examples of model fits in a healthy control subject.** Results of 1T3P, 2T4P, and 2T5P model fittings performed on the lungs, spleen, bone marrow (sacrum and ilium), tonsils, and selected occipital lymph nodes of an example control subject (Sub06) shown for (**A**) the 0–48 h and (**B**) zoomed on the 0–90 min.

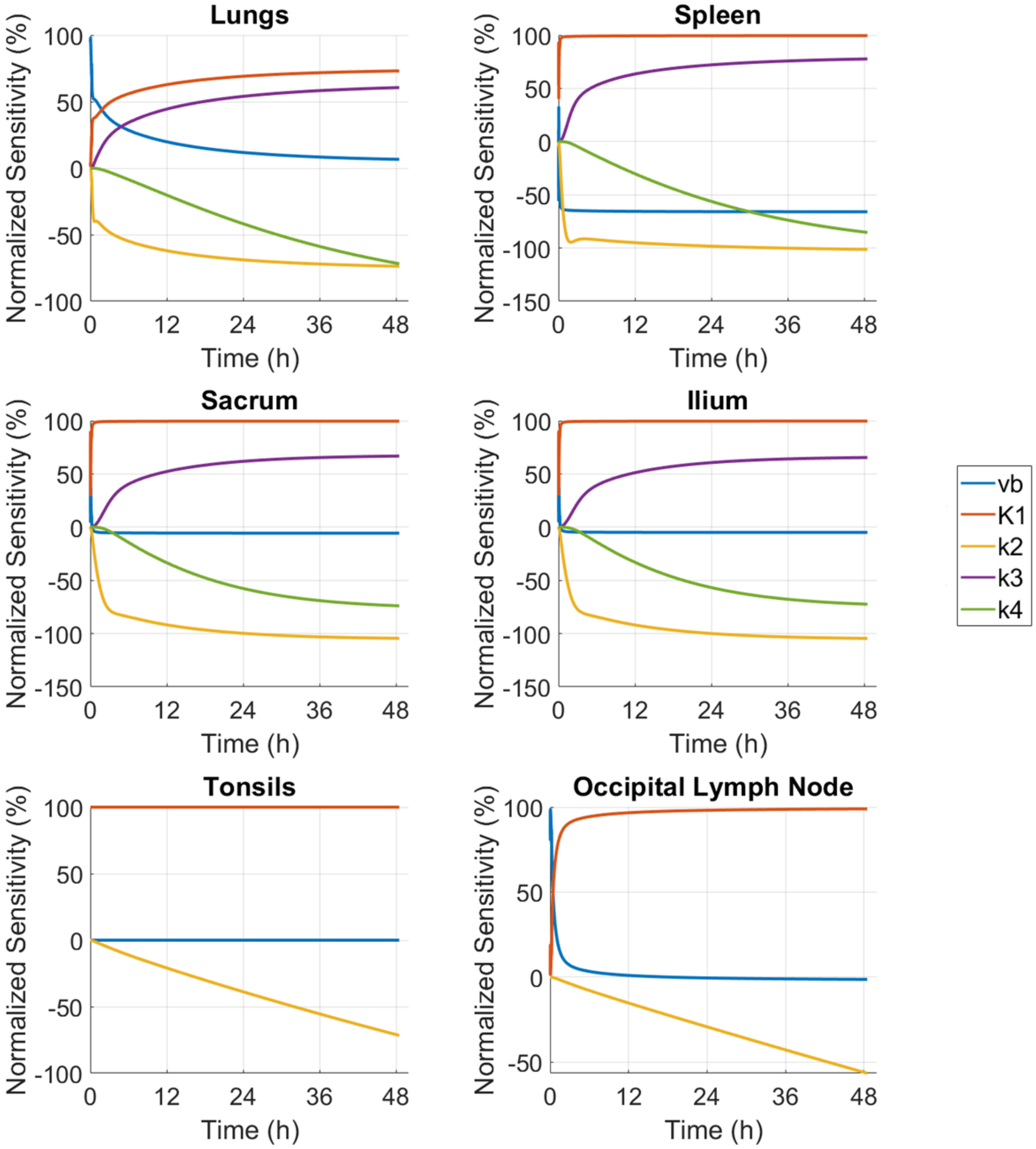


**Fig. S8. Normalized sensitivity plots of microparameters of the AIC-preferred model.** Normalized sensitivities of microparameters of the model with average highest AIC, shown for lungs, spleen, sacrum, ilium, tonsils, and occipital lymph node of the baseline scans of an example COVID-19 patient (Sub02) shown for the 0–48 h.

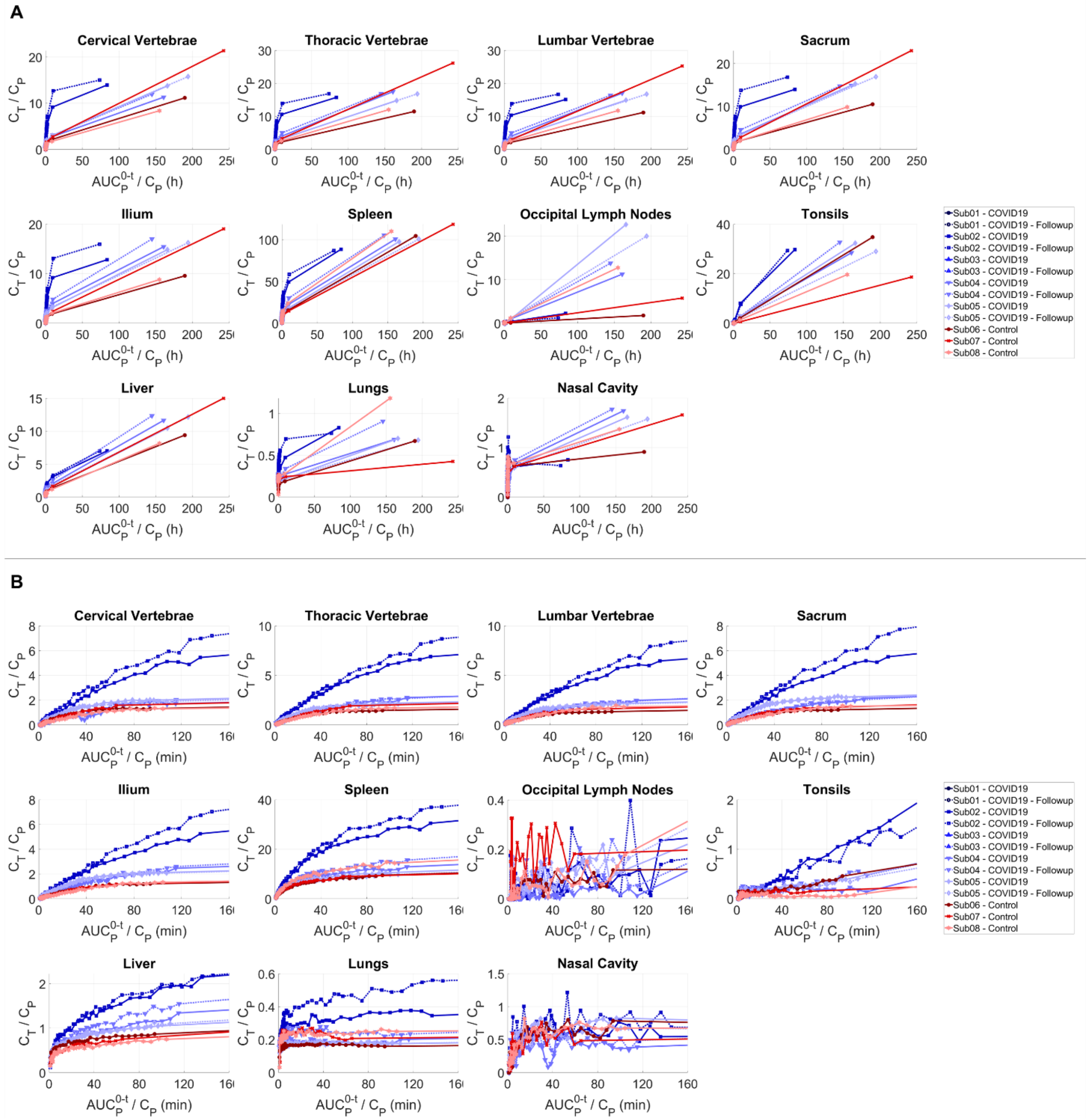


**Fig. S9. Patlak plots of different organs-of-interest.** Patlak plots of different regions of bone marrow, spleen, occipital lymph nodes, tonsils, liver, lungs, and nasal cavity shown (**A**) during the 48-h of the imaging study and (**B**) during the first 90-min after tracer administration for all subjects with dynamic scans.

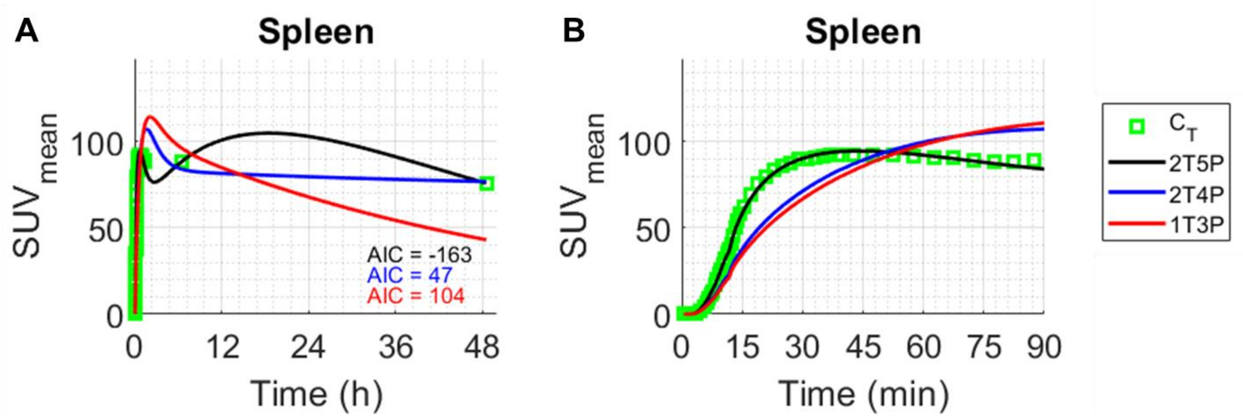


**Fig. S10. Effect of late-timepoint weighting factors on model fitting in spleen.** Results of 1T3P, 2T4P, and 2T5P model fittings performed on spleen of the baseline scans of an example COVID-19 patient (Sub02) shown for (**A**) the 0–48 h and (**B**) zoomed on the 0–90 min, using increased weighting factors (×10 compared to previous fits) for the two late timepoint scans.

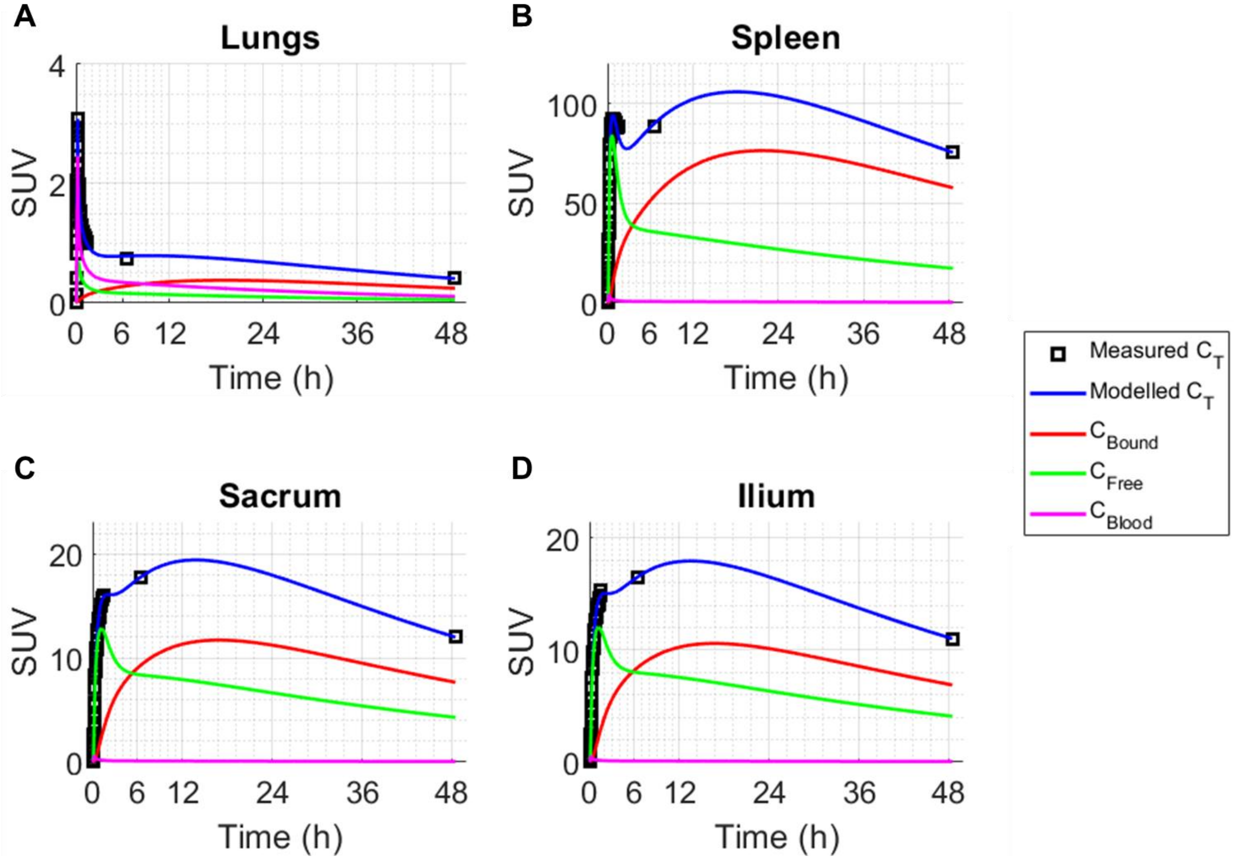


**Fig. S11. Changes in concentrations of 2T5P model compartments as a function of time.** Concentrations of the compartments of the 2T5P model fit performed on (**A**) lungs, (**B**) spleen and bone marrow ((**C**) sacrum and (**D**) ilium) of the baseline scans of an example COVID-19 patient (Sub02) shown for the 0–48 h of the imaging, including the concentrations of free tracer in tissue ($C_{Free}$), bound tracer in tissue ($C_{Bound}$) and the tissue blood fraction ($C_{Blood}$), in which $C_T = C_{Free} + C_{Bound} + C_{Blood}$.

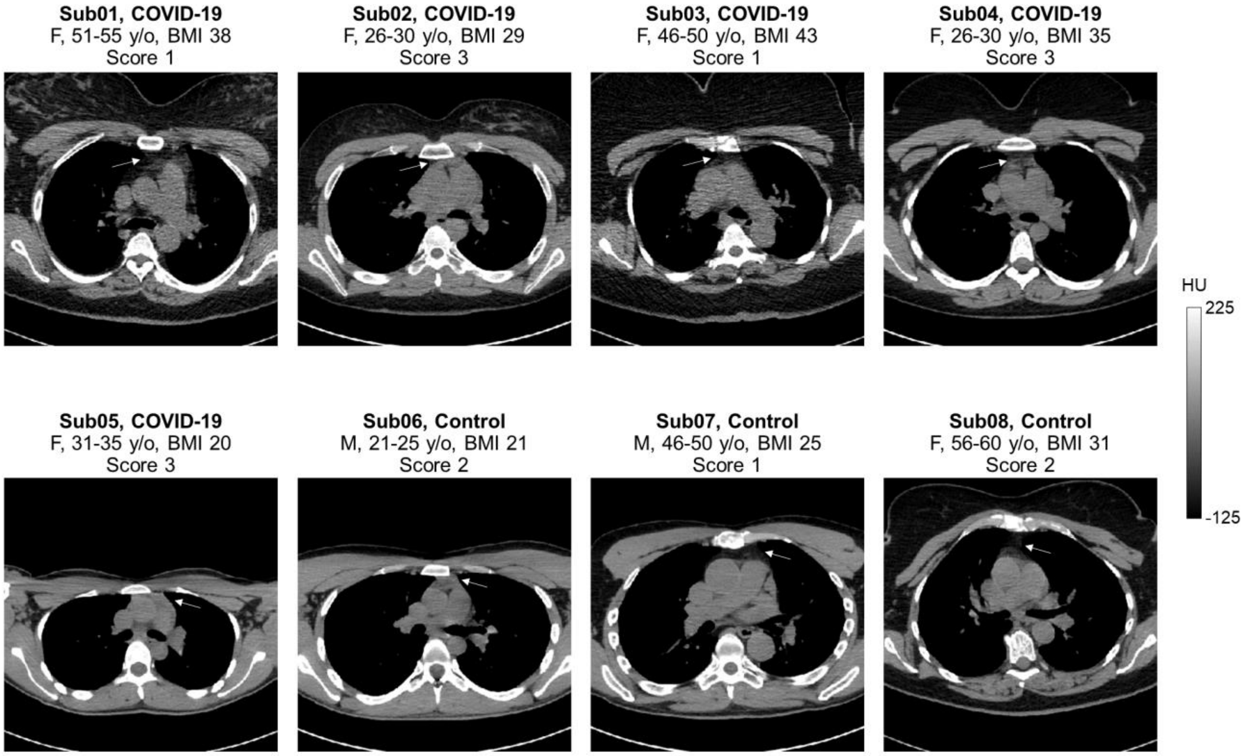


**Fig. S12. CT image slices through the thymus of all subjects.** Selected slices through low-dose CT images of all subjects are compared with the corresponding thymus fatty degeneration scores assigned to each subject.

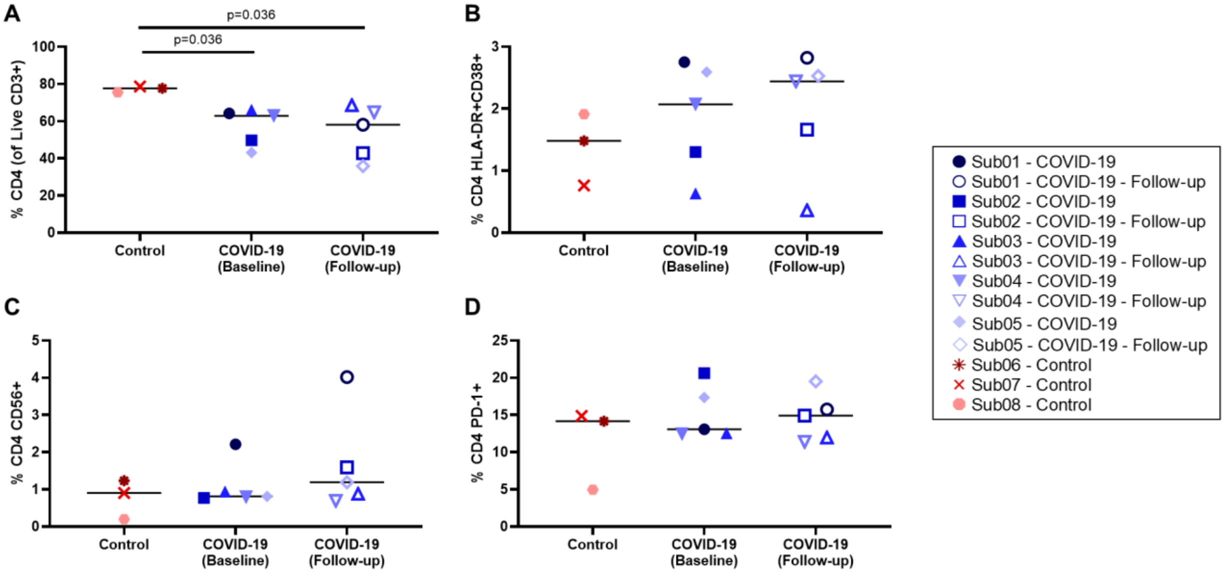


**Fig. S13. Peripheral blood CD4⁺ T cell phenotyping.** (**A**) Percentage of CD4⁺ T cells within the live CD3⁺ population, (**B**) percentage of activated CD4⁺ T cells characterized by CD38 and HLA-

DR co-expression and (**C**) CD56 expression, and (**D**) percentage of exhausted CD4$^+$ T cells characterized by PD-1 expression in all subjects.

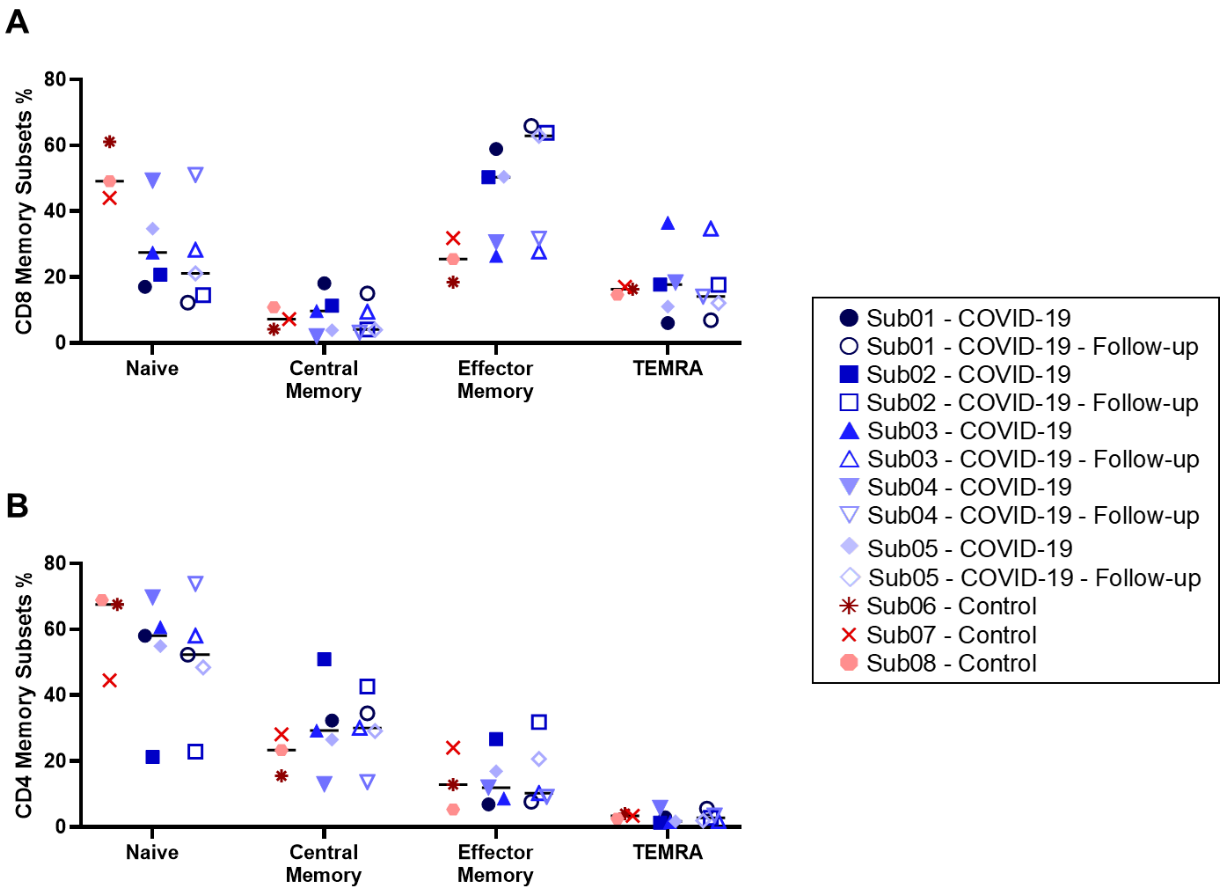


**Fig. S14. Memory subsets of CD8$^+$ and CD4$^+$ T cells.** Memory subsets of (**A**) CD8$^+$ and (**B**) CD4$^+$ T cells, comparing the percentage of naïve, central memory, effector memory, and TEMRA cells in peripheral blood of all subjects.

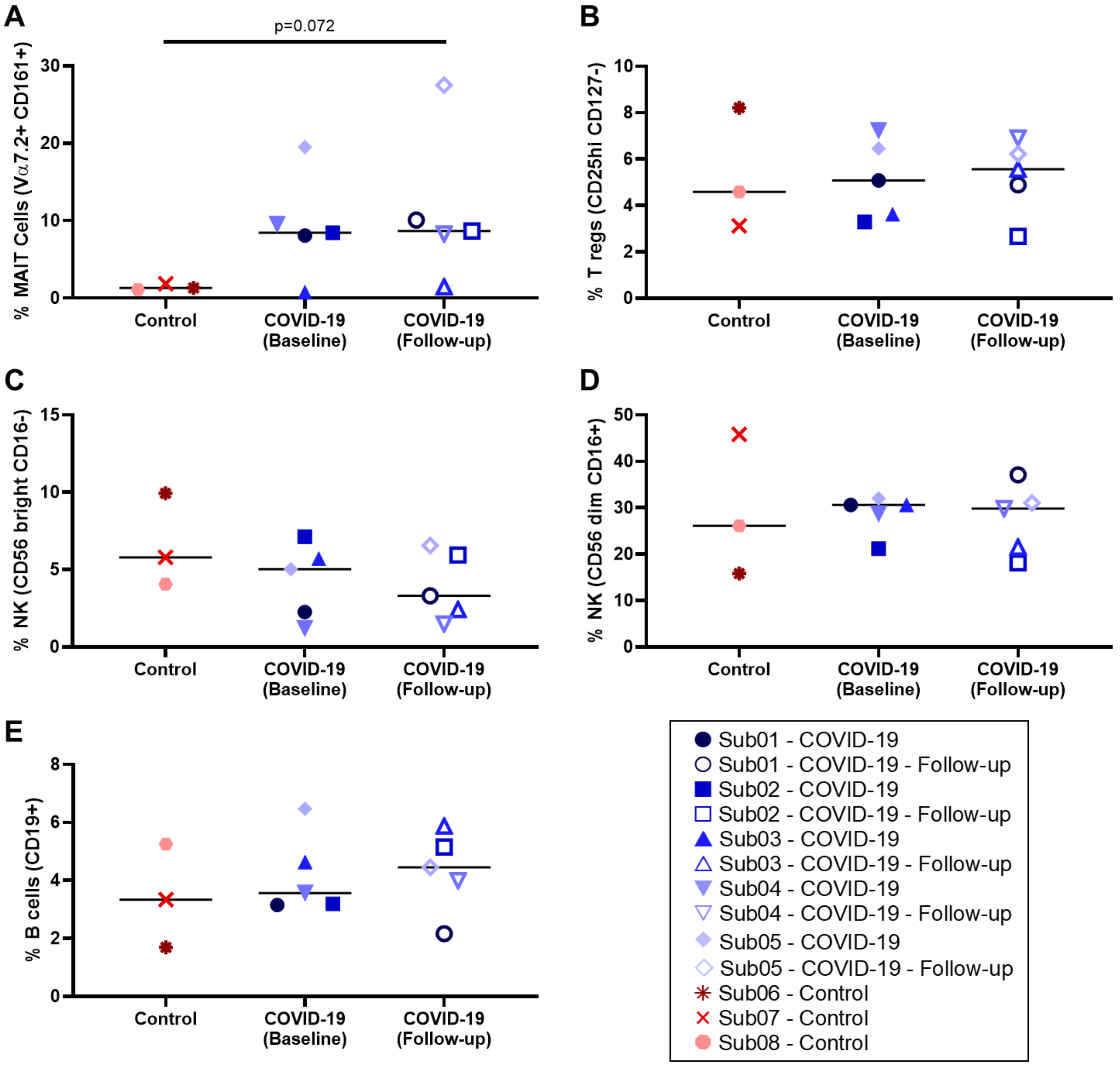


**Fig. S15. Peripheral blood phenotyping of MAIT, $T_{reg}$, NK, and B cells.** Percentages of (**A**) MAIT cells, (**B**) $T_{reg}$ cells, (**C**) cytokine-producing NK cells, (**D**) cytotoxic NK cells, and (**E**) B cells compared in peripheral blood of all subjects.

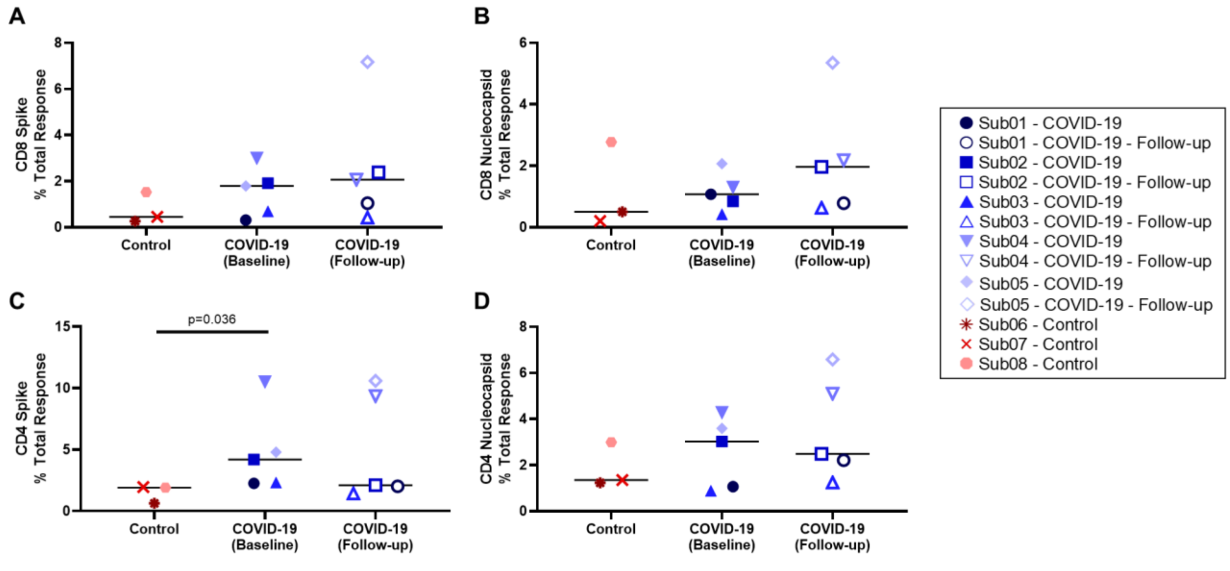


**Fig. S16. Total responses in CD8$^+$ and CD4$^+$ memory T cells.** Total percentage of (**A** and **B**) CD8$^+$ and (**C** and **D**) CD4$^+$ memory T cells responding in any way (CD107a, IFNγ, IL2, MIP-1β, or TNFα) to SARS-CoV-2 (**A** and **C**) spike and (**B** and b) nucleocapsid proteins compared in peripheral blood of all subjects.

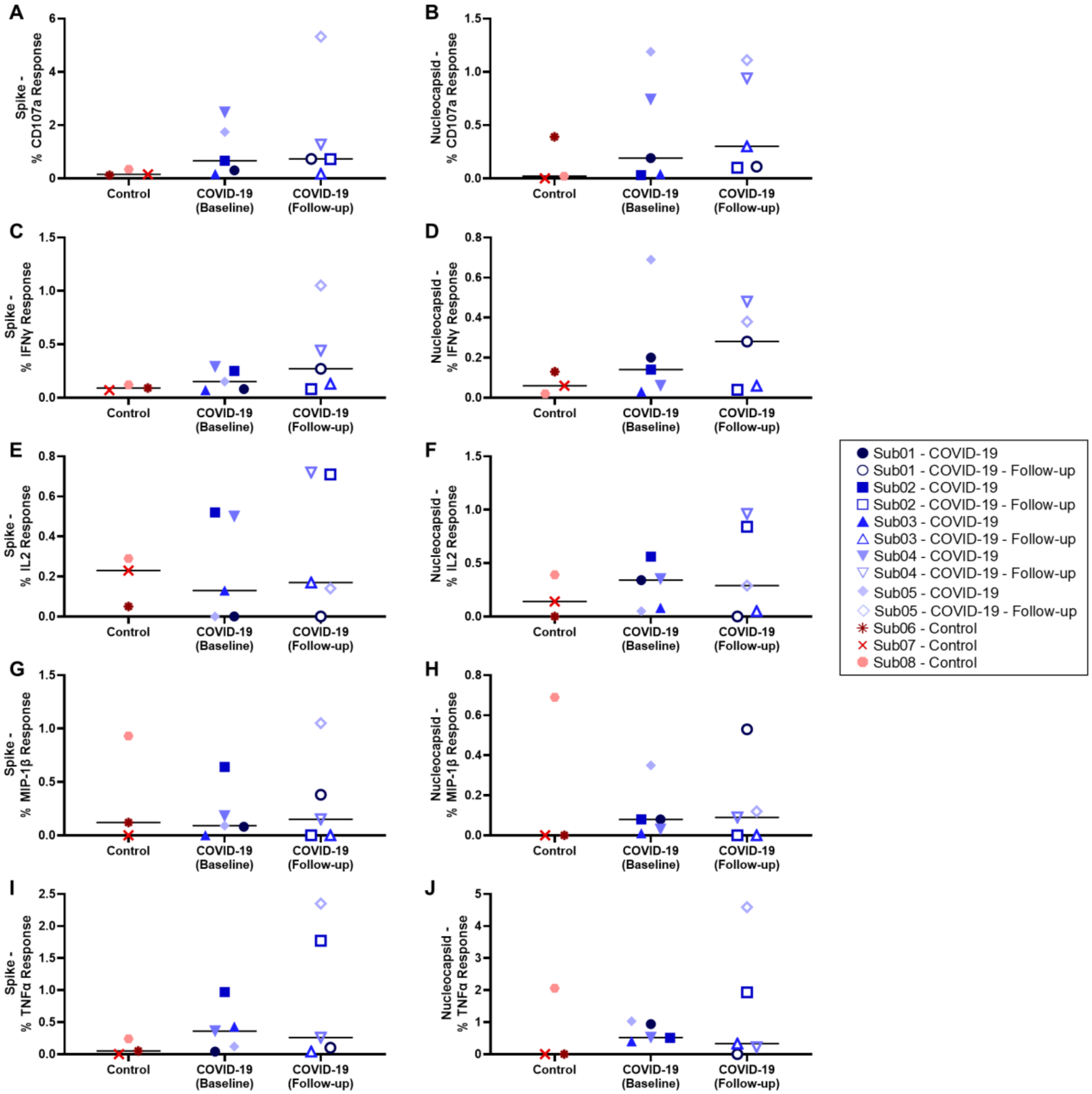


**Fig. S17. Memory CD8$^+$ T cell individual functional responses.** (**A**, **C**, **E**, **G**, **I**) Percentage of SARS-CoV-2 spike-specific and (**B**, **D**, **F**, **H**, **J**) nucleocapsid-specific CD8$^+$ memory T cells showing (**A** and **B**) CD107a expression, (**C** and **D**) IFNγ production, (**E** and **F**) IL2 production, (**G** and **H**) MIP-1β production, and (**I** and **J**) TNFα production.

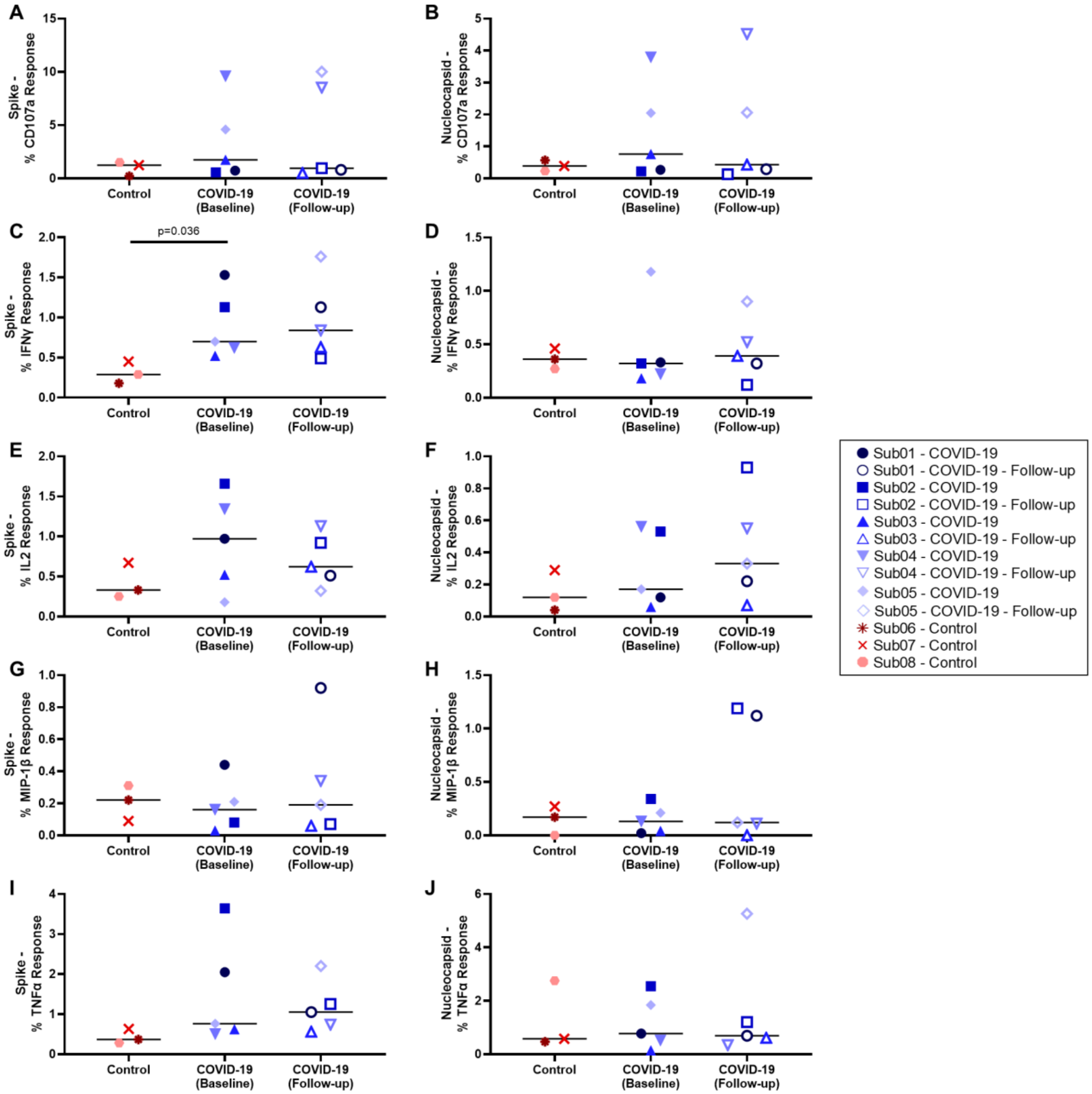


**Fig. S18. Memory CD4+ T cell individual functional responses.** (**A**, **C**, **E**, **G**, **I**) Percentage of SARS-CoV-2 spike-specific and (**B**, **D**, **F**, **H**, **J**) nucleocapsid-specific CD4+ memory T cells showing (**A** and **B**) CD107a expression, (**C** and **D**) IFNγ production, (**E** and **F**) IL2 production, (**G** and **H**) MIP-1β production, and (**I** and **J**) TNFα production.

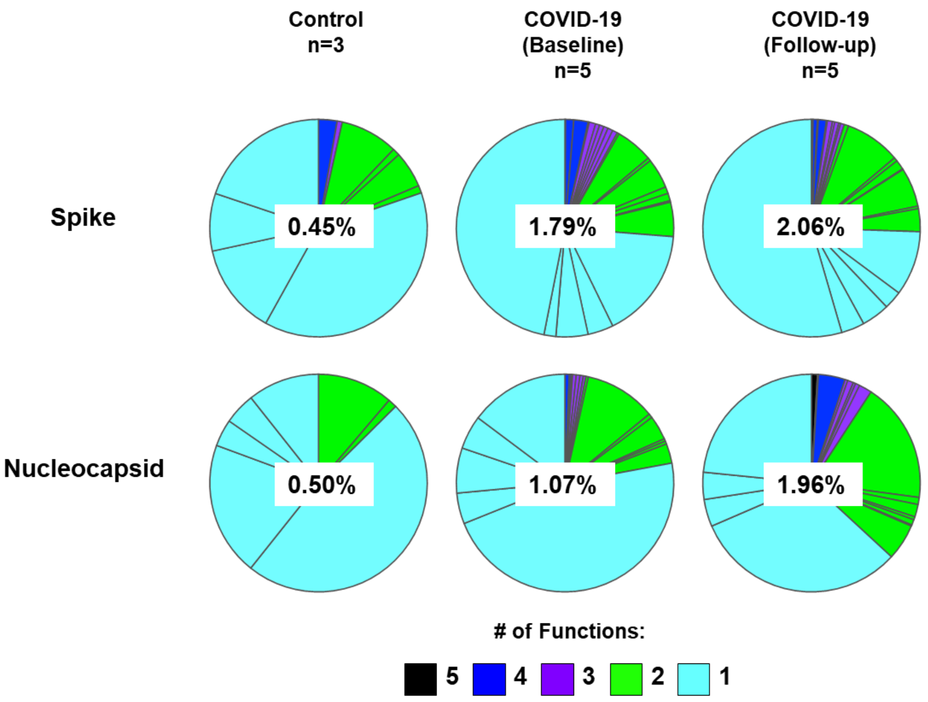


**Fig. S19. Polyfunctional responses in SARS-CoV-2 spike-specific and nucleocapsid-specific CD8$^+$ memory T cells.** The percentages shown on each pie chart represent the median magnitude of the total response in each group.

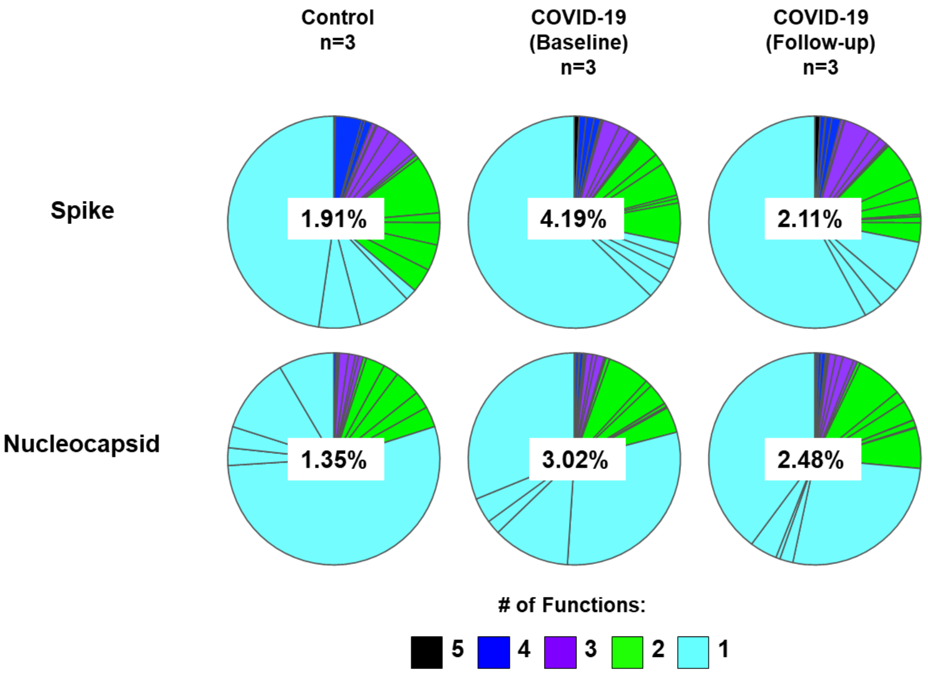


**Fig. S20. Polyfunctional responses in SARS-CoV-2 spike-specific and nucleocapsid-specific CD4$^+$ memory T cells.** The percentages shown on each pie chart represent the median magnitude of the total response in each group.

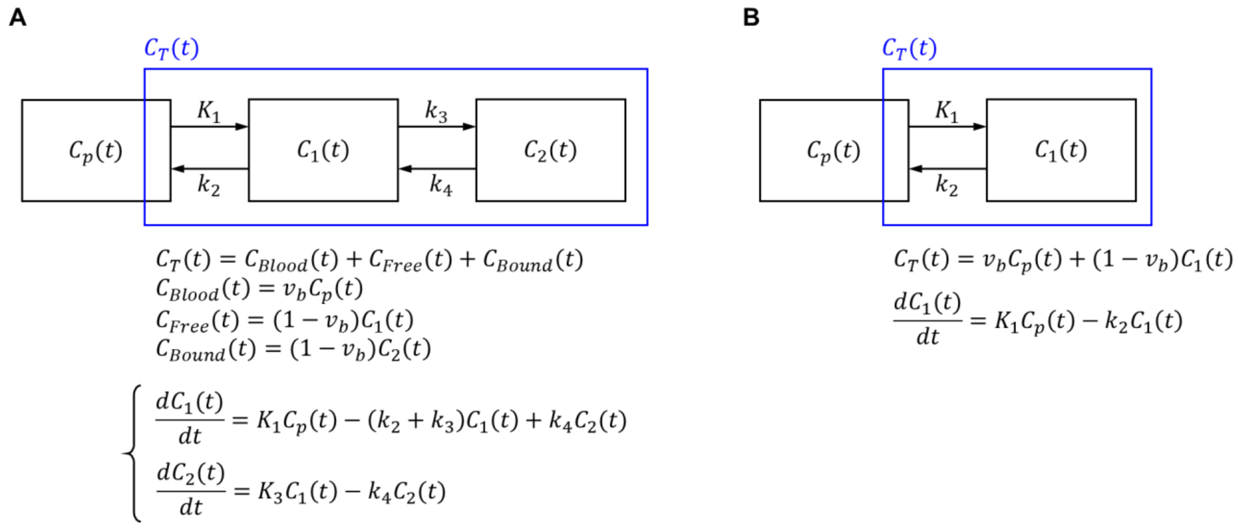


**Fig. S21. Definition of conventional one-tissue and two-tissue compartmental models used in PET kinetic modeling.** (**A**) Two-tissue compartmental model with five fitting microparameters (2T5P) of ($\boldsymbol{v_b, K_1, k_2, k_3, k_4}$) and (**B**) one-tissue compartmental model with three fitting microparameters (1T3P) of ($\boldsymbol{v_b, K_1, k_2}$), in which $\boldsymbol{C_p}$ corresponds to the activity concentration of the tracer in blood plasma (which is approximated by image-derived whole-blood activity concentration in this study), $\boldsymbol{C_T}$ corresponds to the activity concentration of the tracer in tissue, $\boldsymbol{v_b}$ represents the fractional blood volume in tissue, and $\boldsymbol{K_1, k_2, k_3,}$ **and** $\boldsymbol{k_4}$ are the rate constants between the model compartments. Two-tissue compartmental model with fours fitting microparameters (2T4P) can be defined as a special case of 2T5P model, in which $\boldsymbol{k_4}$ is zero. $\boldsymbol{K_1}$ is expressed in mL/min/mL$_{\text{tissue}}$, while $\boldsymbol{k_2, k_3,}$ **and** $\boldsymbol{k_4}$ are all expressed in 1/min. The differential equations defining each model are shown below each model. $\boldsymbol{K_1}$ **and** $\boldsymbol{k_2}$ are affected by the blood flow and the permeability of the tracer across the blood capillaries. In a simplified case, the first compartment represents concentrations of the free tracer in tissue and the second compartment represents concentrations of the tracer bound to cell receptors in the tissue. In the case of 1T3P model, the free and bound compartments are not distinguishable from each other. In the specific case of $^{89}$Zr-Df-Crefmirlimab targeting the CD8 receptors, all rate constants include effects from cell trafficking and the concentrations in each compartment also include concentrations of labeled cells trafficking in and out of the tissue.

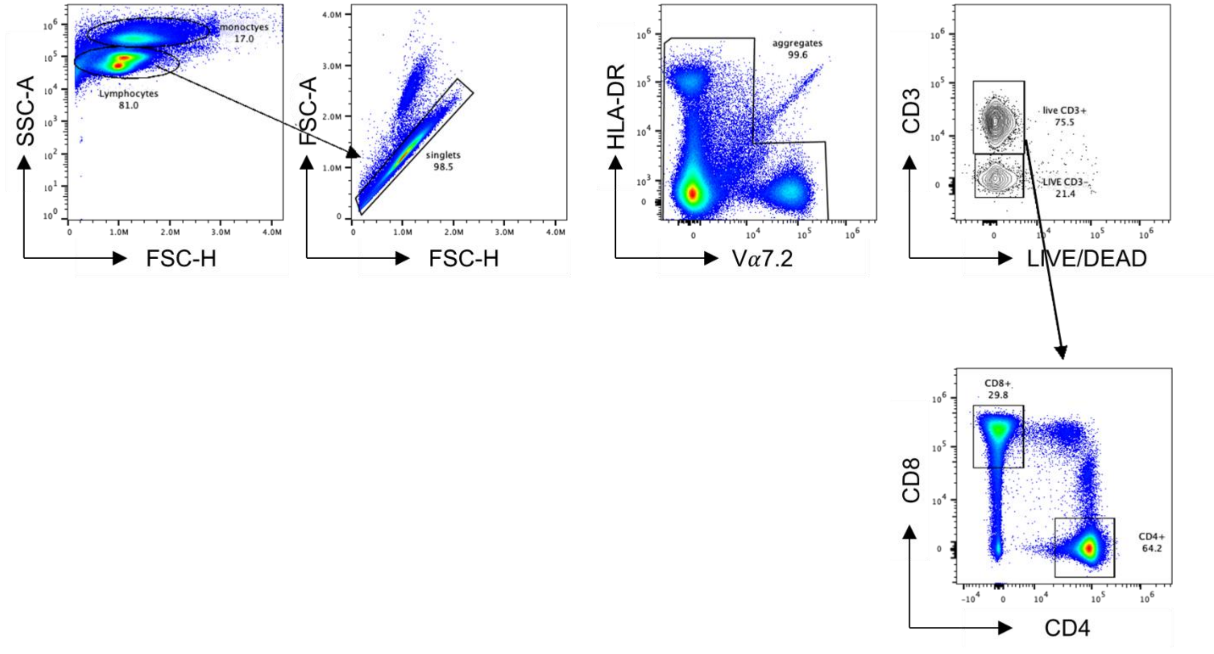


**Fig. S22. Gating strategy for T cell immunophenotyping.** Single cell lymphocytes were gated, aggregates removed, and stained with LIVE/DEAD cell stain kit to exclude non-viable cells. Viable $CD3^+$ cells were gated as T cells and $CD8^+$ and $CD4^+$ T cells were separated.

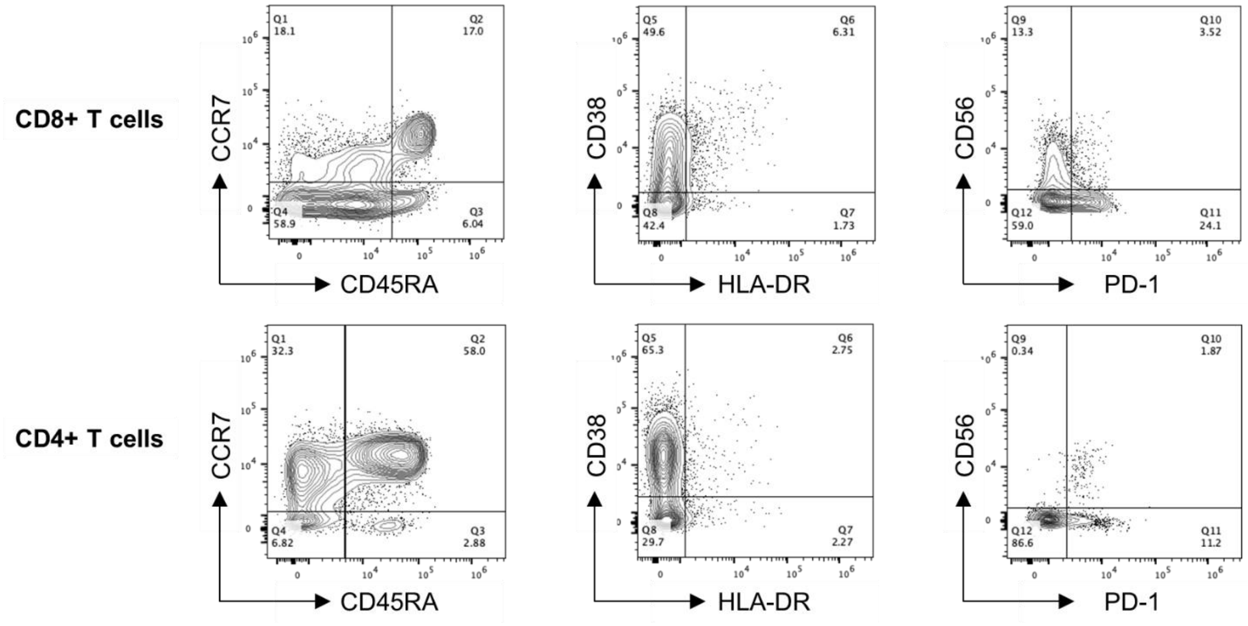


**Fig. S23. Gating strategy for memory T cell immunophenotyping**. CD45RA and CCR7 were used for gating T cell memory subsets, defined as naïve ($CD45RA^+$, $CCR7^+$), central memory ($CD45RA^-$, $CCR7^+$), effector memory ($CD45RA^-$, $CCR7^-$), and terminally differentiated effector memory ($CD45RA^+$, $CCR7^-$). T cell activation was assessed with co-expression of HLA-DR and CD38, in addition to CD56 expression. Lastly, PD-1 expression was used for quantifying the exhausted T cell population.

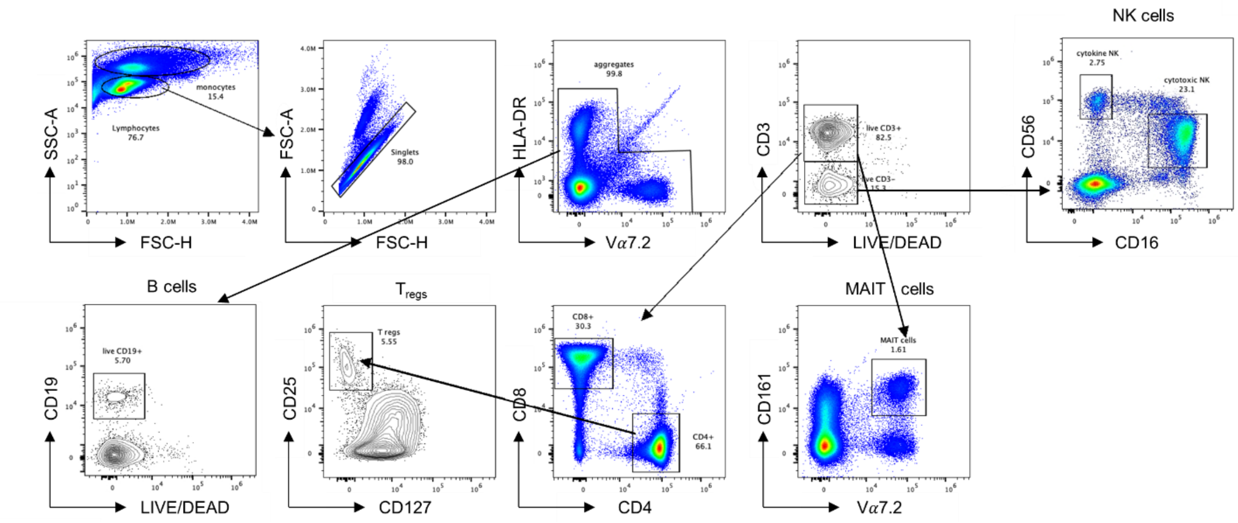


**Fig. S24. Gating strategy for phenotyping T$_{reg}$ cells, MAIT cells, NK cells and B cells.**

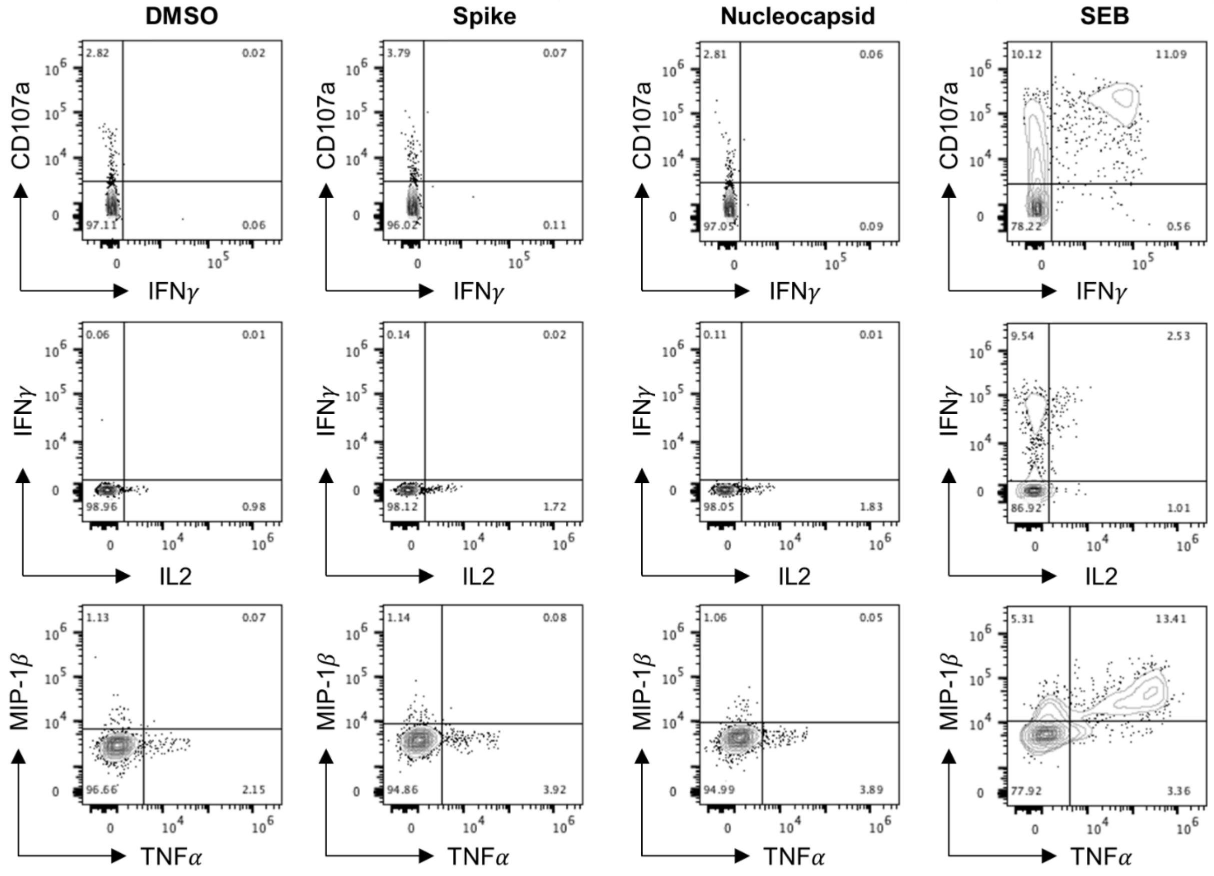


**Fig. S25. Gating strategies to define SARS-CoV-2-specific CD8+ memory T cell response, using individual SARS-CoV-2 peptide pools.** Representative examples of flow cytometry plots of SARS- CoV-2-specific CD8$^{+}$ memory T cells are shown after overnight stimulation with spike and nucleocapsid peptide pools, compared to negative control (DMSO) and positive control stimulation with SEB.
